# Computer-assisted analysis of polysomnographic recordings improves inter-scorer associated agreement and scoring times

**DOI:** 10.1101/2022.03.23.22272801

**Authors:** Diego Alvarez-Estevez, Roselyne M. Rijsman

**Author notes:** **Corresponding author:** Diego Alvarez-Estevez, PhD, Center for Information and Communications Technology Research (CITIC), University of A Coruña, Edif. CITIC, planta 2, Campus Elviña s/n, 15071 A Coruña, Spain.

## Abstract

**Study Objectives:** To investigate inter-scorer agreement and scoring time differences associated with visual and computer-assisted analysis of polysomnographic (PSG) recordings.

**Methods:** A group of 12 expert scorers reviewed 5 PSGs that were independently selected in the context of each of the following tasks: (i) sleep stating, (ii) detection of EEG arousals, (iii) analysis of the respiratory activity, and (iv) identification of leg movements. All scorers independently reviewed the same recordings, hence resulting in 20 scoring exercises from an equal amount of different subjects. The procedure was repeated, separately, using the classical visual manual approach and a computer-assisted (semi-automatic) procedure. Resulting inter-scorer agreement and scoring times were examined and compared among the two methods.

**Results:** Computer-assisted sleep scoring showed a consistent and statistically relevant effect toward less time required for the completion of each of the PSG scoring tasks. Gain factors ranged from 1.26 (EEG arousals) to 2.41 (limb movements). Inter-scorer kappa agreement was also consistently increased with the use of supervised semi-automatic scoring. Specifically, agreement increased from K=0.76 to K=0.80 (sleep stages), K=0.72 to K=0.91 (limb movements), K=0.55 to K=0.66 (respiratory activity), and K=0.58 to K=0.65 (EEG arousals). Inter-scorer agreement on the examined set of diagnostic indices did also show a trend toward higher Interclass Correlation Coefficient scores when using the semi-automatic scoring approach.

**Conclusions:** Computer-assisted analysis can improve inter-scorer agreement and scoring times associated with the review of PSG studies resulting in higher efficiency and overall quality in the diagnosis sleep disorders.

## 1. INTRODUCTION

The analysis of polysomnographic sleep recordings (PSGs) constitutes one of the most time-consuming tasks in the daily work of a Sleep Center. A typical PSG examination contains somewhere between eight up to twenty-four hours of continuous neurophysiological activity recording. Common PSG data include, among others, different traces of electroencephalographic (EEG), electrooculographic (EOG), electrocardiographic (ECG), electromyographic (EMG), and respiratory activity [1]. Likewise, analysis of the PSG can be organized into different subtasks, for instance, analysis of the macro and micro structure of sleep, characterization of the respiratory function, or identification and scoring of limb movement activity.

Clinical findings over the last years have uncovered the negative consequences that Sleep Disorders exert over health, contributing to the general public awareness. This situation has led to a steady increase in the demand for PSG investigations, which represents a challenge for the already congested sleep centers. Clinician’s time is expensive and scant. In addition, the large amount and the complexity of the associated data, makes of PSG analysis a task prone to errors and to subjective interpretations [2]. Indeed, despite homogenization procedures promoted by development and usage of clinical standard guidelines [1] [3], different grades of intra-and inter-expert variability have been reported in the literature, affecting the resulting PSG outcomes, which vary among the specific references or tasks subject to evaluation [4] [5] [6] [7] [8] [9]. In this context, the use of automatic scoring algorithms presents potential advantages. First, as computer analyses produce deterministic (repeatable) outputs, they have the capacity to overcome the variability associated with intrinsic human subjectivity, thereby contributing to standardization of the process and overall quality improvement. On the other hand, automatic scoring would result in great savings in terms of scoring time, hence human resources, reducing overall costs of the diagnosis. Literature, in fact, is rich on examples that focus on the development and validation of automatic analysis methods in different areas related to the scoring of sleep studies [5] [10] [11] [12] [13] [14] [15] [16] [17] [18]. However, despite recent advances and the promising man-machine agreement results reported in some of these works, reliance on automatic scoring among the clinical community remains low [19] [20] [21]. Moreover, there are still open questions on whether “are we there yet”, in terms of acceptable performance and enough generalization capabilities of these algorithms as compared to well-trained human clinicians [2].

An alternative approach is the so-called “semi-automatic” scoring, where an automatic algorithm performs a preliminary analysis pass, after which the clinician reviews its results editing possible miss-scorings. Because of the expert supervision, this procedure avoids the extended concern that (full) automatic scoring would fail in the presence of data of varying quality and/or different patient phenotypes. Semi-automatic scoring, therefore, effectively results in valid outcomes both in the context of research and routine clinical practice. It remains an open question, though, whether economics of semi-automatic scoring still apply, in particular, because of keeping (at least partial) manual review as part of the process, and its possible role in reducing the baseline levels of human scoring variability.

The main goal of this study is to evaluate the possible benefits of using semi-automatic analysis for the scoring of PSGs in comparison to baseline levels of human performance. Performance, in the context of this study, is characterized by the use of objective (quantifiable) metrics regarding the respective scoring times and resulting levels of inter-rater scoring variability. Specifically, in this study, experiments are carried out, independently and systematically, in the context of the following four (usually, the most important) PSG analysis subtasks: sleep staging, analysis of EEG micro-arousals, evaluation of the respiratory function, and scoring of limb movements.

Our work is of interest, as the topic has barely been examined in the available literature, with perhaps some few exceptions regarding the specific subtask of sleep staging [19] [22] [23]. Regardless, to our knowledge, no previous studies have attempted to examine the hypothesis on whether semi-automatic scoring can contribute to reduction of inter-scorer variability in a systematic way. Likewise, we believe this is the first study to systematically address possible scoring time differences between the manual and semi-automatic approaches.

To have objective (measurable) references of the levels of scoring performance is as well of fundamental importance to allow implementation of quality control mechanisms in the patient care. Our work contributes as well by adding to the existing literature on manual scoring, taking into account that evolution of the scoring methods and reference clinical guidelines motivates reassessment of the existing references, for which some of them might be outdated. Furthermore, for some of the examined scoring tasks, literature references of related quality metrics examined in this work have never been reported before.

## 2. METHODS

### 2.1. Study database

PSG data for this study has been gathered by retrospective inspection of the Haaglanden Medisch Centrum (HMC, The Hague, The Netherlands) Sleep Center clinical patient database. All data were acquired in the course of the common clinical practice. No patient was therefore subjected to any additional behavior in relation to this study, nor was prescribed any additional treatment outside of the regular clinical workflow.

Recordings were de-identified and subrogate study numbers were assigned to each patient, avoiding any possibility of individual patient identification. Under these conditions the study obtained approval of the local Medical Ethics Committee (Medisch Ethische Toestsingscomissie Zuidwest Holland) under code MTEC-19-065, who considered that the protocol did not fall under the scope of the Medical Scientific Research Involving Human Subjects Act (WMO) and that no explicit informed consent was required by participants. Study has as well obtained written permission from the database owner for publication.

PSG data consisted of raw biomedical signals following standard acquisition procedures according to the AASM guidelines [1]. SOMNOscreen™ plus devices (SOMNOmedics, Germany) were used as the acquisition hardware. Event annotations resulting from the scorers’ reviews during regular clinical workflow accompanied each recording. Clinical scorings were carried out by HMC sleep technicians including the scoring of sleep stages, EEG arousals, and respiratory activity, following the AASM standards [1], and the scoring of the limb movement activity following the WASM2016 guidelines [3]. Both the raw signal’s data and the resulting clinical scoring annotations were digitally stored using the EDF+ format [24].

### 2.2. Rescoring task

In the present study a group of sleep technicians were prompted to review 5 PSGs that were independently selected in the context of each of the following tasks: (i) sleep stating, (ii) detection of EEG arousals, (iii) analysis of the respiratory activity, and (iv) identification of leg movements. Hence, in total, 20 different PSG recordings were included in the study, were on each case, scoring was limited to the specific task under consideration. All sleep scorers belonged to the same sleep center (HMC) and have demonstrated experience participating regularly, and autonomously, in the scoring of PSGs in the regular clinical workflow. Sleep technicians undergoing training or supervision were excluded from this study. In total, 12 scorers participated in this task.

Each of the participant scorers was tasked to review the exact same recordings. In all cases, scorers were blinded to both the patient identity (by using de-identified recordings) and possible results of previous scorings (e.g. that took place during regular clinical workflow, from other scorers, or during a previous self-rescoring subtask).

Rescoring was repeated, separately for each task, using first a purely manual, followed by a semi-automatic scoring approach, hence resulting in a total 40 different scoring exercises per scorer. To avoid learning effects, at least 4 months of separation were scheduled between these two scoring moments. For reference, the average amount of 70 PSG recordings are scored by each sleep technician due to the normal sleep lab activity during that period. Scorers were also not informed of the fact that manual and semi-automatic scorings would involve the exact same recordings.

All scoring tasks took place using the Polyman software [25]. For each task, a timer was automatically set in the background by the program (unavailable to the human scorer). The tick counting was automatically paused if no mouse or keyboard interaction was detected during more than a minute, and the offline time was subtracted from the total scoring time. The resulting active scoring time periods were saved separately in a file for later analysis.

Scoring took place between Time In Bed periods only (between lights off – on markers), which were provided as pre-filled annotations. For scoring of EEG arousals, respiratory events and limb movements, the pre-filled clinical hypnogram was also provided as additional source for contextual interpretation. Scorers were instructed to stick to the scoring of the relevant events in the context of the specific target task, not being allowed to change any pre-filled contextual information, if provided.

During the semi-automatic scoring process, the annotations that resulted from the output of the corresponding automatic analysis algorithms were provided, in addition, at the start of the scoring. Scorers were instructed to review these scorings by adding, deleting, or editing the event’s onset and offset times, where corresponds, and according to their own expertise. Details regarding the development and validation of the automatic scoring algorithms that were used for this purpose have been reported in past works. The reader is referred to check the corresponding references regarding the automatic scoring of sleep stages [2] [26], leg movement activity [27] [28], respiratory events [29] [30], and EEG arousals [31] [32].

### 2.3. Selection of PSGs

Each of the 5 PSGs selected for each scoring task (i.e. sleep staging, EEG arousals, respiration, and limb movements) were selected from an initial pre-sample dataset of 2801 recordings, corresponding to the most recent one-year data from the HMC database at the time (2019). Sampling size per task was determined by limited resources with regard to expert time available for the study. As result, 20 PSG recordings were selected that integrate the final scoring sample for the study.

The selection process was implemented with the objective to minimize the chance of selection bias and obtain a balanced representation of scoring difficulty for each task. The hypothesis is that scoring difficulty is correlated with scoring time and inter-scorer variability: the more difficult is to score a PSG by an expert scorer, the more time it would take and the more inter-scorer variability would be associated to its scoring, and viceversa.

No specific exclusion criteria were applied during the selection process to filter out recordings, for instance, due to specific patient conditions, or poor signal quality. A sufficient condition was that the recording had been accepted for manual scoring during regular clinical workflow, a condition that, by definition, was already satisfied by all recordings included in the pre-sample dataset. The underlying motivation was to reproduce, as close as possible, the same conditions as in real practice, by considering a representative sample of the general patient phenotype.

Hence, for each of the four scoring subtasks, the following selection procedure was scheduled in which human-automatic agreement is used as subrogate of scoring difficulty:

i. Taking as reference the complete pre-sample dataset (2801 PSGs) a full automatic analysis (no human intervention at all) of the recording was performed. This analysis led to a list of automatically scored events *L*_*a*_*(i)*, for each recording *i*, related to the corresponding scoring task under consideration.
ii. Using the list of automatically-generated events, *L*_*a*_*(i)*, each PSG was compared with the corresponding list of events that resulted during routine clinical examination, *L*_*c*_*(i)*. Confronting *L*_*a*_*(i)* with *L*_*c*_*(i)*, a preliminary metric of performance agreement between the two scoring outputs, *K*_*ac*_*(i)*, was obtained. Specifically, *K*_*ac*_ was calculated using the Cohen’s Kappa statistic [33]. Details on the implementation of *K*_*ac*_ for each of the four target subtasks are described in the “analysis methods” section.
iii. By repeating this operation through all 2801 PSGs available in the initial pre-sample dataset, a distribution *DK*_*ac*_ of *K*_*ac*_*(i)* values was obtained.
iv. Using *DK*_*ac*_ as reference, uniform sampling was performed to select the target number *N=5* of recordings to be included in each subtasks’ final study dataset. Specifically, the 5 recordings whose associated *K*_*ac*_*(i)* performance metrics represent the middle of each inter-quartile range, plus the median, were selected as representatives of their respective whole populations. In other words, the recordings with performance scores representing the 12.5th, 37.5th, 50th, 62.5th and 87.5th percentiles of each *DK*_*ac*_ distribution were selected for the final study dataset.

Effectively, the above described procedure is preferable over random resampling as it avoids potential selection of outliers by chance (i.e. extreme favorable or unfavorable cases for the automatic algorithm) that might bias the resulting sample. Similar selection procedures were scheduled during the validation of different automatic scoring algorithms that were reported in the past [32] [26].

Correlation analyses are scheduled to analyze validity of the selection hypothesis whose results are provided in Supplementary Table D1 and discussed in the corresponding Section D of the Supplementary Materials.

Table 1 summarizes the general demographics and PSG descriptors in the resulting patient study sample. Data are presented stratified among the corresponding task-specific subgroups.

**Table 1.** Summary of general demographics and PSG descriptors in the study dataset. PSG descriptors correspond to values resulting from retrospective examination in the clinical database, i.e. prior to the multi-expert rescoring procedures carried out in this study. Distributions are characterized using the median and the corresponding interquartile ranges

| Parameter | Group values |  |  |  |  |
| --- | --- | --- | --- | --- | --- |
|  | Subset A | Subset B | Subset C | Subset D | Total |
| n | 5 | 5 | 5 | 5 | 20 |
| Age (years) | 52.0 [47.0, 57.0] | 57.0 [51.0, 68.0] | 59.0 [57.0, 61.0] | 55.0 [52.0, 63.0] | 57.0 [51.8, 61.5] |
| Male (n, %) | 5 (100%) | 1 (20%) | 3 (60%) | 3 (60%) | 12 (60%) |
| Time In Bed (TIB, hours) | 7.5 [7.4, 8.0] | 7.3 [7.0, 7.3] | 6.5 [6.4, 7.4] | 8.1 [7.2, 8.2] | 7.3 [7.0, 8.0] |
| Total Sleep Time (TST, hours) | 5.9 [5.9, 7.1] | 6.7 [5.9, 6.7] | 6.0 [5.8, 6.1] | 7.3 [5.5, 7.4] | 6.0 [5.7, 7.0] |
| Sleep Latency (SL, min) | 4.6 [3.0, 5.4] | 1.8 [1.5, 3.3] | 9.7 [2.6, 14.5] | 1.9 [1.0, 21.5] | 3.1 [1.6, 16.2] |
| Stage R latency (min) | 69.5 [40.0, 174.0] | 104 [64.5, 124.0] | 59.0 [52.0, 138.0] | 83.5 [80.0, 85.0] | 81.8 [58.0, 127.0] |
| Wake After Sleep Onset (WASO, min) | 72.8 [24.7, 124.0] | 53.0 [39.9, 68.2] | 38.9 [28.1, 52.2] | 89.6 [46.7, 100.4] | 52.6 [27.3, 106.3] |
| Sleep Efficiency (SE, %) | 91.1 [74.2, 94.5] | 88.6 [83.8, 90.9] | 89.9 [84.7, 92.8] | 83.0 [76.6, 90.5] | 89.2 [76.0, 93.2] |
| Arousal Index (Arl, n/TST) | 4.7 [1.8, 17.6] | 14.3 [13.4, 16.0] | 19.8 [9.6, 23.4] | 9.6 [7.9, 13.7] | 13.5 [7.5, 20.7] |
| Apnea-Hypopnea Index (AHI, n/TST) | 9.1 [6.0, 9.4] | 5.1 [0.6, 20.7] | 6.2 [5.8, 13.1] | 7.2 [3.0, 20.6] | 6.7 [5.0, 20.6] |
| Oxygen Desaturation Index (ODI, n/TST) | 10.7 [10.5, 10.8] | 1.4 [0.6, 24.8] | 9.9 [5.5, 15.4] | 9.9 [2.8, 15.7] | 10.2 [4.4, 18.0] |
| Leg Movement Index (n/TST) | 52.3 [9.0, 56.5] | 32.0 [21.0, 57.7] | 14.2 [8.6, 39.8] | 30.0 [19.3, 43.2] | 31.0 [12.9, 53.4] |
| Periodic Leg Movement Index (PLMI, n/TST) | 13.0 [0.3, 45.7] | 13.8 [13.4, 49.0] | 0.8 [0.4, 26.7] | 22.4 [4.1, 28.4] | 13.6 [0.7, 32.8] |

Subset-scoring task correspondences: A - sleep stages; B - Limb Movements; C - Respiratory Events; D - EEG arousals

### 2.4. Analysis methods

Analysis of inter-scorer agreement is carried out in the first place by discretizing the recording time into non-overlapping analysis mini-epochs. Each analysis mini-epoch is assigned the corresponding scorer’s output in the context of the specific target subtask. Duration of the mini-epochs are task-related as well. In the case of the sleep scoring, analysis epochs have the standard duration of 30s and take possible values according to the AASM clinical guidelines, that is, either W, N1, N2, N3, or R [1]. In the context of the EEG arousals, respiratory events, and limb movement’s scoring subtasks, each mini-epoch takes a binary value noting the presence or absence of event, respectively, if overlapping or not with the events marked by the scorer. Analysis mini-epoch duration is set to 0.5s for all the three subtasks.

Time discretization in the above terms leads to the construction of *k*-dimensional contingency tables (*k*=5 for sleep staging, *k*=2 otherwise) from which standard metrics of agreement for categorical data can be derived. Within each task, agreement between each of the twelve scorers’ pair combination (n = 66 per recording) is calculated using the Cohen’s kappa statistic. The use of the Cohen’s kappa is motivated given its widespread use in the field, as well as its robustness in the case of imbalanced class distributions as it corrects for agreement due to chance [33].

Inter-scorer agreement is also evaluated among the diagnostic indices resulting from the respective scorings. Following the list of recommended parameters to be reported in PSG studies [1] [3], a representative subset for each of the subtasks targeted in this study is selected. In particular, sleep quality-related parameters of Sleep Efficiency (SE), Sleep Onset Latency (SOL), and Wake After Sleep Onset (WASO) [34], in relation to the sleep scoring task; Apnea-Hypopnea Index (AHI), Apnea Index (AI), Hypopnea Index (HI) and Oxygen Desaturation Index (ODI), in relation to the scoring of respiratory events; Arousal Index (ArI), in relation to the scoring of EEG arousal events; and Leg Movement Index (LMI) and the Periodic Leg Movement Index (PLMI), in relation to the limb movement scoring task. LMI and PLMI indices are calculated according to the WASM2016 scoring guidelines, the former being defined as the number of limb movements ≥ 0.5s after bilateral combinations per hour of sleep, with the latter including respiratory-related LMs as well in the counts [3]. Inter-scorer agreement among the resulting indices on each case (n = 12 per recording) is evaluated using the Intraclass Correlation Coefficient (ICC) [35]. Specifically, a two-way absolute single-measures variant of the statistic, ICC(A,1), is used [36]. A Matlab implementation for calculation of this coefficient has been used whose source code is available at [37].

Hypothesis testing is carried out to check for significant differences between the manual and semi-automatic scoring approaches. For this purpose, the reference level for statistical significance is set to α = 0.05. Differences are examined using the paired version of the Wilcoxon signed rank test among all the matched kappa scorer pair combinations (n = 66 per recording, n = 330 in total for each task). Analogous analysis is performed for checking out differences in the respective scoring times among the matched individual scorers (n = 12 per recording, n = 60 in total for each task). For each test the corresponding effect size is reported using the Cohen’s *D* statistic. Statistical significance on inter-scorer ICC agreement differences among diagnostic indices is also evaluated. For this purpose, the a priori expected agreement (*r0*) for the semi-automatic approach is set to the effective ICC levels achieved with manual scoring.

Results of the above-mentioned analyses are presented in the subsequent section by aggregating the respective scorings among the five recordings involved within each scoring task. In order to keep the main text extension attainable, individualized per-recording results are provided as Supplementary Material. In this case, manual vs. semi-automatic differences in diagnostic indices are examined, again, using paired analyses. Comparison of the respective variance distributions is examined using the Brown-Forsythe (unpaired) test. For the latter, i.e. comparison of distribution’s variance, the corresponding manual and semi-automatic indices are first mean normalized within their respective distributions to avoid possible bias due to differences in the respective population means.

## 3. RESULTS

### 3.1. Analysis of scoring time

Figure 1 shows the median scoring time associated with the completion of the different analysis tasks according to the followed approach, i.e. manual or semi-automatic. Values on the bar plot are shown in minutes and aggregate the results among the five recordings involved on each case.

**Figure 1.**
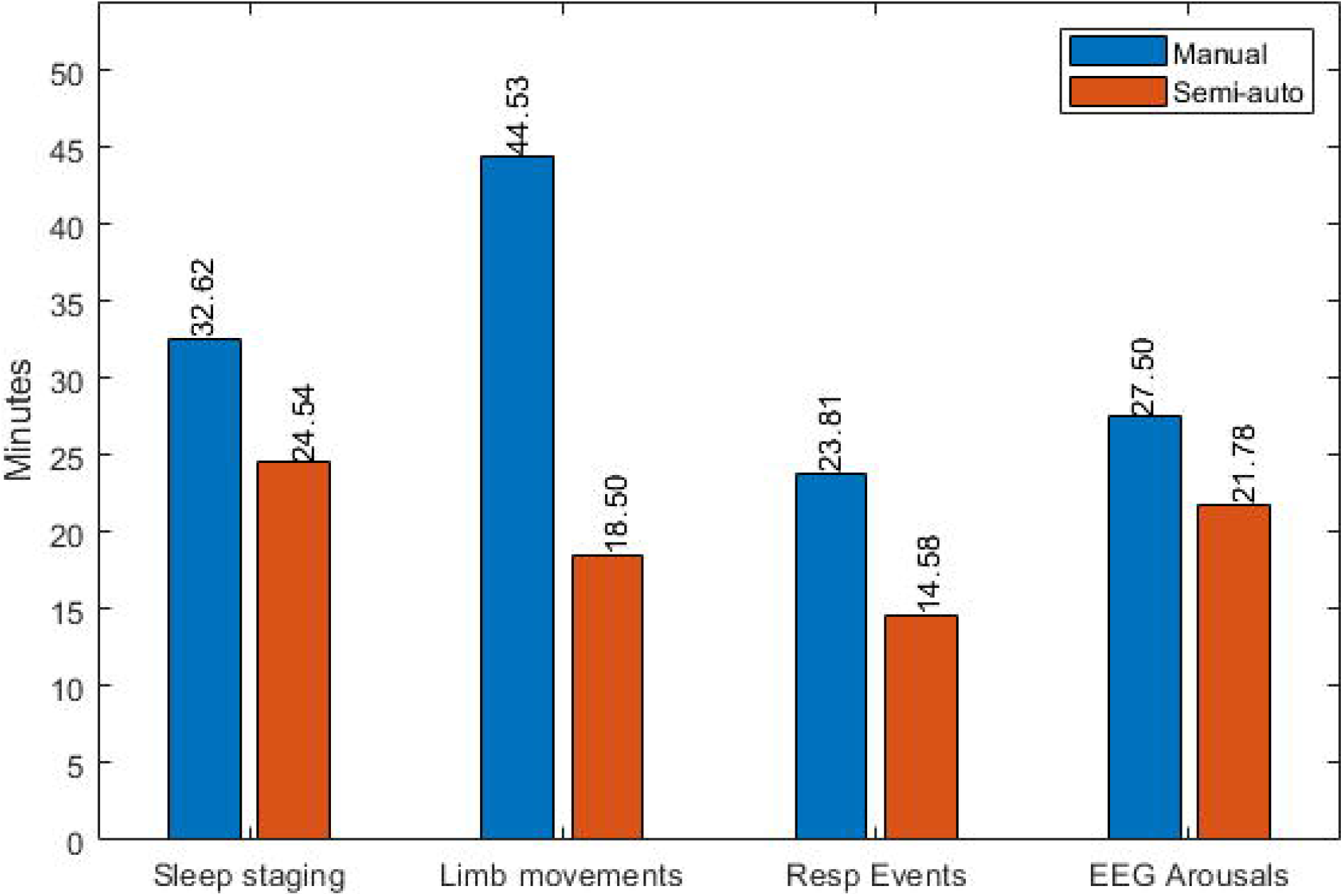
Differences in scoring time between manual and semi-automatic scoring approaches. Median scoring time per task is shown in minutes.

Table 2 expands the results of

**Table 2.**
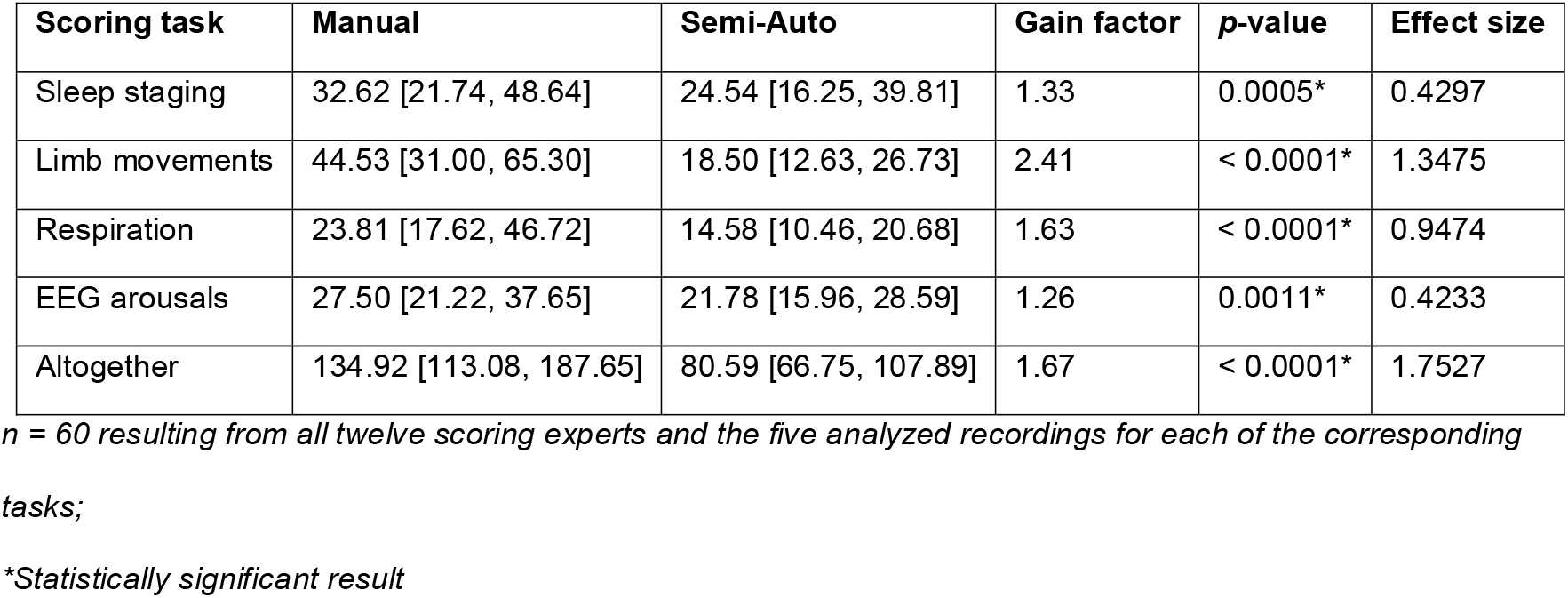
Analysis of scoring time differences per task between manual and semi-automatic approaches. Distributions are characterized using the corresponding median and interquartile ranges in minutes. Gain factors are calculated on each case as the ratio between the corresponding median scoring times

| Scoring task | Manual | Semi-Auto | Gain factor | p-value | Effect size |
| --- | --- | --- | --- | --- | --- |
| Sleep staging | 32.62 [21.74, 48.64] | 24.54 [16.25, 39.81] | 1.33 | 0.0005* | 0.4297 |
| Limb movements | 44.53 [31.00, 65.30] | 18.50 [12.63, 26.73] | 2.41 | < 0.0001* | 1.3475 |
| Respiration | 23.81 [17.62, 46.72] | 14.58 [10.46, 20.68] | 1.63 | < 0.0001* | 0.9474 |
| EEG arousals | 27.50 [21.22, 37.65] | 21.78 [15.96, 28.59] | 1.26 | 0.0011* | 0.4233 |
| Altogether | 134.92 [113.08, 187.65] | 80.59 [66.75, 107.89] | 1.67 | < 0.0001* | 1.7527 |

*n = 60 resulting from all twelve scoring experts and the five analyzed recordings for each of the corresponding tasks;*
*\*Statistically significant result*

Figure 1 and shows the results of the associated statistical analyses involving the two scoring approaches. Data in Table 2 unveil a consistent and statistically relevant effect toward less time required for the completion of each task when using the semi-automatic scoring approach. Gain factors vary per task, with the largest time savings relating to the scoring of limb movements, followed by the analysis of the respiratory activity, and a less pronounced effect associated with the sleep staging task and the scoring of EEG arousals. The associated effect sizes on each case support these interpretations. In this regard, notice that a positive sign on the corresponding index indicates that the overall effect (in this case scoring time) is bigger in the manual scoring scenario, with the associated absolute value being an indicative of how much bigger the effect is.

When comparing absolute time values among the different tasks, our results show that identification of limb movements is the most time consuming task when using manual analysis. Scoring of respiratory events is relatively the quickest. The trend changes a bit when using the semi-automatic approach, resulting in sleep staging being the slowest, with analysis of respiratory activity remains as the fastest task.

Individualized per-recording and per-scorer analyses for each task can be found, respectively, in Supplementary Tables A1-A4 and Supplementary Figures A1-A4.

### 3.2. Analysis of kappa agreement

Figure 2 shows the global kappa agreement results per scoring task when comparing manual and semi-automatic scoring approaches. Values on the bar plot represent the median expert paired agreements among the five recordings within the corresponding task.

**Figure 2.**
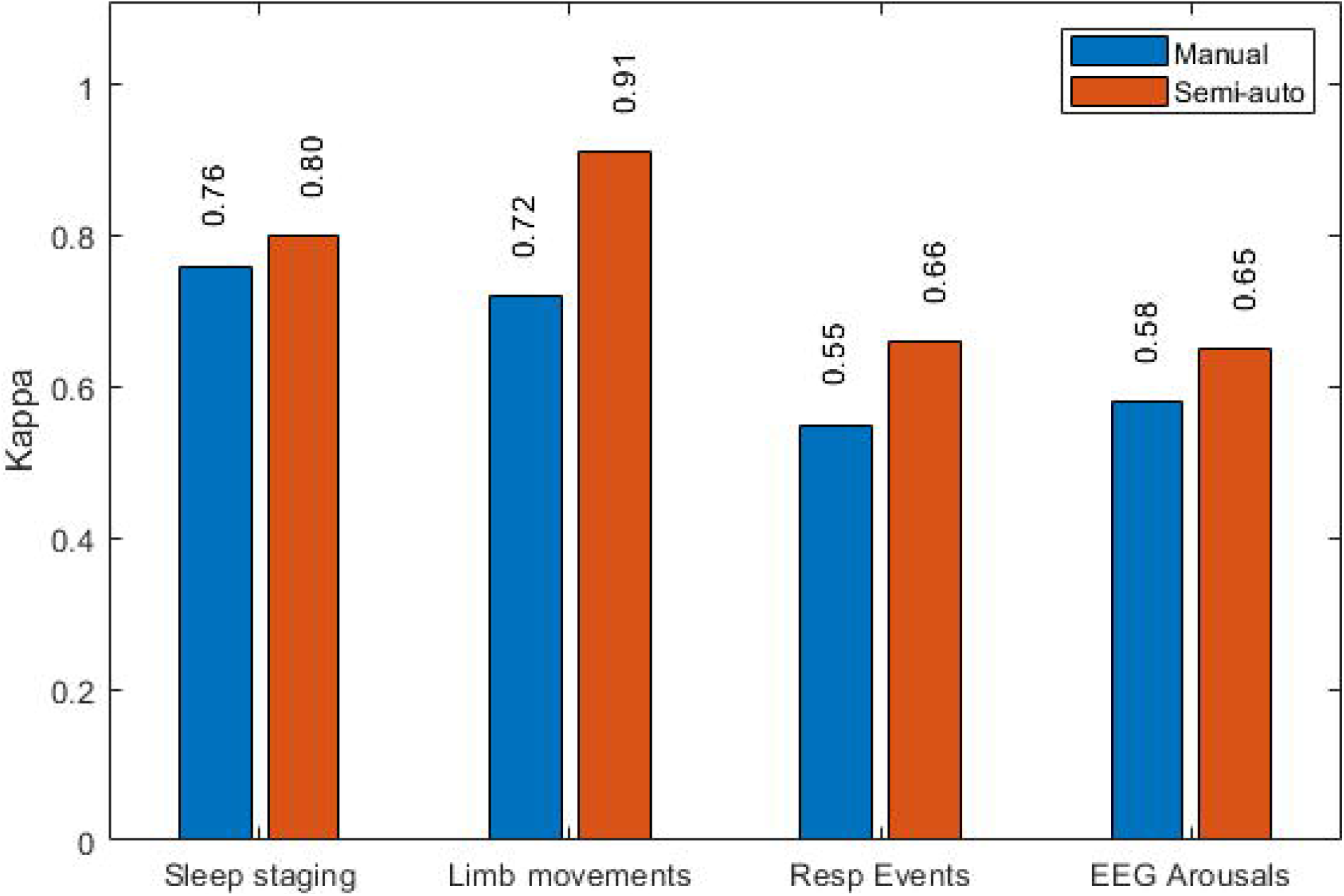
Agreement comparison between manual and semi-automatic scoring approaches. Median Kappa agreement for each task is shown by aggregating all the expert pair combinations throughout the five corresponding recordings.

Table 3 shows results of the statistical analyses between the corresponding manual and semi-automatic scoring differences. Moreover, results are subcategorized for some of the tasks into different contexts of clinical interest. In particular, differences between wake and sleep periods are reported for limb movements, as well as results involving different types of events within the respiratory analysis task. For the analysis of the limb movement activity, the individual kappa scores for each leg channel (left / right) were averaged together before statistical analysis was executed.

**Table 3.** Overall kappa inter-scorer agreement per scoring task and comparison between manual and semi-automatic approaches. Distributions are characterized using the corresponding median and interquartile ranges. TIB = Time in Bed

| Scoring task | Context | Manual | Semi-auto | p-value | Effect size |
| --- | --- | --- | --- | --- | --- |
| Sleep staging | TIB | 0.76 [0.69, 0.80] | 0.80 [0.76, 0.83] | < 0.0001* | -0.7146 |
| Limb Movements | TIB | 0.72 [0.64, 0.79] | 0.91 [0.86, 0.95] | < 0.0001* | -2.0223 |
|  | Wake | 0.67 [0.57, 0.77] | 0.89 [0.82, 0.94] | < 0.0001* | -1.7121 |
|  | Sleep | 0.75 [0.65, 0.81] | 0.92 [0.86, 0.95] | < 0.0001* | -1.6704 |
| Respiratory activity | Apnea, Hypopnea, RERA (TIB) | 0.55 [0.43, 0.78] | 0.66 [0.53, 0.89] | < 0.0001* | -0.8315 |
|  | Apneas (TIB) | 0.74 [0.35, 0.88] | 0.88 [0.57, 0.98] | < 0.0001* | -0.3783 |
|  | Hypopneas (TIB) | 0.46 [0.36, 0.53] | 0.61 [0.51, 0.68] | < 0.0001* | -0.9569 |
| EEG Arousals | TIB | 0.58 [0.48, 0.65] | 0.65 [0.56, 0.71] | < 0.0001* | -0.6166 |

*n* = 330 resulting from all distinct combinations of expert scorer pairs (*n* = 66) on each of the five recordings involved in the corresponding scoring task;
\*Statistically significant result

Results from Table 3 show that statistically significant differences between manual and semi-automatic scoring are reached regardless of the specific task or the event subtype. A consistent trend toward higher inter-scorer agreement associated with the use of semi-automatic scoring is shown. Notice the associated effect sizes overall show a negative sign, being indicative of the general smaller agreement achieved in the manual scoring scenario. The highest absolute effect in this context is associated with the analysis of the limb movement activity task.

When comparing among the different tasks, the highest (either manual or semi-automatic) agreements are achieved in the case of the sleep staging and limb movements’ analysis tasks. For the latter, higher agreement is obtained during sleep periods than during wakefulness. With regard to the analysis of respiratory activity, and attending to the different event subtypes, higher agreement is achieved for the scoring of apneas than of hypopneas. Finally, reliability associated with the scoring of EEG arousal events reaches agreement levels similar to those obtained for the identification of respiratory events in general (i.e. apneas, hypopneas and RERAs altogether).

Individualized per-recording analyses for each of the tasks are supplied in Supplementary Tables B1-B8.

### 3.3. Analysis of derived diagnostic indices

Table 4 examines inter-scorer agreement among the selected list of diagnostic parameters for the manual and semi-automatic scoring approaches. Agreement is evaluated using the Interclass Correlation Coefficient (ICC).

**Table 4.** Comparison of inter-scoring agreement among diagnostic indices between manual and semi-automatic approaches. Agreement is characterized in terms of Interclass Correlation Coefficient (ICC), and corresponding index distributions using the respective median and interquartile ranges. r0 = null hypothesis on ICC score; SE = Sleep Efficiency; SOL = Sleep Onset Latency; WASO = Wake After Sleep Onset; (P)LMI = (Periodic) Leg Movement Index; AHI = Apnea-Hypopnea Index; AI = Apnea Index; HI = Hypopnea Index; ODI = Oxygen Desaturation Index; ArI = Arousal Index.

| Index (TST) | Summary of index distributions | | ICC | | $r_0$ | ICC $p$-value | |
| --- | --- | --- | --- | --- | --- | --- | --- |
|  | Manual | Semi-auto | Manual | Semi-auto |  | Manual | Semi-auto |
| SE (%) | 89.11 [68.05, 94.87] | 90.31 [70.59, 94.86] | 0.9938 | 0.9974 | 0 | < 0.0001* | < 0.0001* |
|  |  |  |  |  | 0.9938 | --- | 0.0665 |
| SOL (min) | 5.27 [2.50, 107.41] | 4.11 [2.31, 104.81] | 0.8720 | 0.8367 | 0 | < 0.0001* | < 0.0001* |
|  |  |  |  |  | 0.8720 | --- | 0.5589 |
| WASO (min) | 86.20 [22.27, 151.77] | 79.18 [23.02, 139.51] | 0.9924 | 0.9970 | 0 | < 0.0001* | < 0.0001* |
|  |  |  |  |  | 0.9924 | --- | 0.0481* |
| LMI | 25.27 [19.78, 70.19] | 31.73 [20.89, 61.82] | 0.9227 | 0.9757 | 0 | < 0.0001* | < 0.0001* |
|  |  |  |  |  | 0.9227 | --- | 0.0166* |
| PLMI | 13.91 [9.20, 61.70] | 14.14 [12.79, 53.70] | 0.9351 | 0.9771 | 0 | < 0.0001* | < 0.0001* |
|  |  |  |  |  | 0.9351 | --- | 0.0282* |
| AHI | 5.86 [3.71, 12.47] | 6.54 [4.70, 14.61] | 0.9878 | 0.9924 | 0 | < 0.0001* | < 0.0001* |
|  |  |  |  |  | 0.9878 | --- | 0.1901 |
| AI | 1.26 [0.31, 3.46] | 1.57 [0.00, 3.54] | 0.9962 | 0.9986 | 0 | < 0.0001* | < 0.0001* |
|  |  |  |  |  | 0.9962 | --- | 0.0441* |
| HI | 3.10 [2.38, 4.87] | 4.55 [3.13, 6.68] | 0.6010 | 0.7540 | 0 | < 0.0001* | < 0.0001* |
|  |  |  |  |  | 0.6010 | --- | 0.1160 |
| ODI | 6.48 [4.24, 14.38] | 12.04 [6.01, 15.89] | 0.8370 | 0.9843 | 0 | < 0.0001* | < 0.0001* |
|  |  |  |  |  | 0.8370 | --- | < 0.0001* |
| Ari | 17.57 [12.98, 25.64] | 18.38 [13.82, 25.41] | 0.6824 | 0.7673 | 0 | < 0.0001* | < 0.0001* |
|  |  |  |  |  | 0.6824 | --- | 0.2290 |

*n = 60 resulting from all twelve scoring experts and the five analyzed recordings for each of the corresponding tasks;*
*\*Statistically significant result*

When comparing absolute ICC values among the different tasks, a trend can be seen toward higher inter-scorer agreement when using the semi-automatic scoring approach, with the only exception of SOL. Regardless of the scoring approach, the highest general agreement is achieved in the case of SE, AI, and WASO (ICC > 0.99 in all cases). Agreement associated with the scoring of apneas probably contributes to the relative high scores achieved for the AHI too. Detection of hypopneas as reflected by HI, on the other hand, shows relative lower levels of ICC agreement. HI is, in fact, is the index where the lowest overall agreement is achieved, followed by ArI. All the obtained ICC values, also regardless of the scoring approach, achieve statistical significance when the null hypothesis assumes no a priori agreement (*r0* = 0)

For examining statistical significance of the observed differences between the manual and semi-automatic approaches, the null hypothesis is set, in addition, to assume baseline ICC levels corresponding to manual scoring. In this case, significant differences are obtained for the indices of WASO, LMI, PLMI, AI and ODI. In the case of SE, AHI, HI and ArI, the trend remains consistent toward higher ICC values when using the semi-automatic scoring approach, albeit analyses do not reach statistical relevance. And finally, only for SOL, higher ICC values are obtained using manual scoring, but again differences do not reach the level of statistical significance.

Individualized per-recording analyses are supplied in Supplementary Tables C1-C10. In this case manual vs. semi-automatic differences are examined both using paired Wilcoxon sign-rank and unpaired Brown-Forsythe tests, as described in the methods section.

## 4. DISCUSSION

The main goal of this study was to evaluate the possible benefits of using semi-automatic scoring of PSGs in comparison to classical manual visual approach. For this purpose, we have individually considered four of the most common subtasks involved in the analysis of PSGs: sleep staging, identification of EEG arousals, evaluation of the respiratory function, and scoring of limb movements. On each case, quantifiable metrics of performance regarding the scoring time, and inter-scorer agreement, have been examined and compared among the two methods. To our knowledge, this is the first study to systematically address the differences between manual and semi-automatic scoring.

Our experimentation has shown that the use of semi-automatic analysis has benefits in the form of faster scoring and higher inter-scorer agreement. Faster scoring means reduced associated costs, and the possibility to reduce the waiting lists by the consequent increase in the scoring production. Higher inter-scorer agreement means better consistency and reliability of the PSG outcomes, and therefore improved quality of the diagnosis. The trend is consistent though all of the four examined tasks. Differences between the two approaches have achieved statistical significance both for the scoring time and the expert agreement measured in terms of Cohen’s K. The impact of these differences on a subset of derived diagnostic indices, analyzed in terms of ICC agreement, has shown a more heterogeneous pattern. While statistical significant differences have been observed for indices of WASO, LMI, LMI, AI and ODI, the same was not the case for indices of SE, SOL, AHI, HI and ArI. Still, for all the examined indices with the exception of SOL, the trend was consistent toward higher ICC values when using the semi-automatic scoring approach.

Our study provides a number of firsts. To the authors knowledge, for example, this is the first study reporting and comparing the time associated with the scoring of respiratory events (both for the manual and semi-automatic approaches). Our data shows a median gain factor of 1.63 when using semi-automatic scoring. That we know of, and excluding preliminary estimations from our own group [38] [32], this is also the first study to report on scoring time associated to manual and semi-automatic scoring of limb movements and EEG arousals. Specifically, a 2.41 gain factor (44.53 minutes for the manual approach, and 18.50 minutes when using the semi-automatic procedure) for the limb movement detection task was obtained, similar to that reported previously in Roessen et al. [38], but using an older version of the clinical scoring guidelines. With regard to the scoring of EEG arousals, a gain factor of 1.26 (median of 27.50 minutes for manual vs. 21.78 for semi-automatic) was obtained in the present study, not far from results reported in [32], using the same clinical reference and automatic scoring algorithm, but on a different selection of PSG recordings. Last, with respect to sleep staging, some few works can be found that have examined scoring times associated with this task using different settings. Koupparis et al. [22], for example, have reported an average 3 hours for manual scoring, which could be improved to 45 minutes with the use of semi-automatic scoring with associated inter-scorer agreement of K = 0.61. Baseline agreement for manual scoring, though, was not reported in their study. Younes et al. [23] have shown differences between full and minimal human intervention associated to semi-automatic sleep staging involving 50 and 6 minutes, respectively, on average. Baseline time for manual scoring, however, was not reported in the study. Our data has shown that 1.33 gain factor in scoring time associated to sleep staging is possible when using semi-automatic scoring, in addition, while improving median inter-scorer agreement from K = 0.76 to K = 0.80 at the same time. This is not a trivial result, as some authors have suggested that full-editing of automatic sleep staging would result in about the same time-scoring as manual scoring [19]. A general warning when comparing results from different works in the literature, is that one has always to bear in mind that relevant differences might exist between the respective population samples, the analysis methods, or the clinical scoring references valid at the time of the study.

When regarding analysis of inter-scorer reliability in the scoring of sleep studies literature is more abundant. Here most of data available regard almost exclusively to manual scoring of sleep stages, with only few examples addressing the case of other scoring tasks. We were not able to find any previous references regarding analysis of inter-scorer reliability under the context of semi-automatic scoring.

For the case of manual sleep staging, review of the related literature can be found in another recent publication by the authors [26], resulting in kappa coefficients ranging widely between K = 0.46 - 0.89, depending on the consulted reference. This range is compatible with the median agreement achieved in this study (K = 0. 76 for manual). We were able to find some studies examining inter-scorer agreement of sleep stages in the context of semi-automatic analysis. In Younes et al. [23] paired kappa between two scorers was increased from 0.71 to 0.95 with the use of semi-automatic scoring and the help of a third scorer. In Koupparis et al. inter-scorer agreement reached a maximum average of K = 0.61 with semi-automated analysis of the so-called hypnospectrogram [22]. Similarly, in Svetnik et al. [19] epoch-by-epoch agreement between scorers performing full or partial review of automated scoring ranged K = 0.60 – 0.63. Reference agreement baseline for manual scoring, however, was not reported in the latter works. In our study we have reported an increase from K = 0.76 to K = 0.80 with the use of semi-automatic scoring.

Only one past reference was found examining inter-scorer manual agreement in the detection of limb movements. In the study of Pittman et al. [5] K = 0.77 was obtained between two scoring experts on a dataset of 31 PSGs. Notice, however, that agreement reported in Pittman et al. refers only to the scoring of PLMS, not LMs, and that the scoring reference was based on older standards (ASDA1993 [39]). Moreover, analysis was constrained to sleep periods only, and its associated resolution was 30s. Our results, involving twelve scorers, and using the more recent WASM2016 scoring standards, resulted in a global K = 0.72 for manual, and K = 0.91 for semi-automatic scoring, when examining LMs during TIB, and using a 0.5s analysis step. Agreement falls respectively to K = 0.67 and K = 0.89 during wake periods, and improves to K = 0.75 and K = 0.92 during TST.

Pitman et al. have reported as well a K = 0.82 for the manual scoring of apneas and hypopneas using the 2001 AASM Medicare scoring definitions on a 30s analysis epoch [5]. With our settings, we have achieved rather lower agreement resulting in median K = 0.55 (improving to K = 0.66 with semi-automatic scoring). We have obtained higher agreement for the scoring of apneas (median K = 0.74 for manual, K = 0.88 for semi-automatic) than for the case of hypopneas (respectively K = 0.46 and K = 0.61). This is an expected result, however no study that we know of had attempted to quantify this difference in terms of kappa agreement so far.

As for the EEG arousal task, some studies can be found reporting kappa values for manual scoring in the K = 0.47 – 0.59 range [32] [40] [41]. Once again, some of these studies use older scoring guidelines (ASDA1992) besides other sources of variability, and therefore direct comparison has to be carefully considered. Regardless, the reported range is consistent with our experimental results in the case of manual scoring (K = 0.58). Our study shows, in addition, that better inter-scorer agreement can be achieved if using semi-automatic scoring (up to K = 0.65 in our dataset).

In this study we have also examined possible differences among the resulting derived indices for clinical diagnosis. Here again, literature regarding inter-rater variability in the context of semi-automatic scoring is very limited. Svetnik et al. have reported no significant differences between manual and semi-automatic scoring in resulting indices for SOL and WASO [19]. This matches our results in the case of SOL, but not for WASO. Koupparis et al., on the other hand, have reported ICC values for WASO of 0.91 and SE of 0.93 under full-editing semi-automatic review, decreasing considerably to ICC of 0.05 and 0.20, respectively, under minimal editing [22]. In our case, both values achieved ICC around 0.99. We were not able to find any other references in the literature for the remaining indices examined in our study.

In the context of manual analysis, on the other hand, one can find several other past studies reporting on inter-rater related ICC agreement scores [5] [42] [7] [43] [44] [45] [6] [46] [47]. The specific values of agreement vary per study. Danker-Hopfe et al. [6] and Kuna et al. [47], for example, have reported ICC values for SE of 0.91 and 0.77 respectively, which is below the agreement obtained in this study for manual scoring (ICC = 0.99). Reliability on PLMS has been reported by Pittman et al. [5] (ICC = 0.93) and Bliwise et al. [42] (ICC = 0.91 – 0.99), however, using older definitions of the index [48] [49]. This is relevant as recent studies [50] [51] [52] have pointed out to significant differences in the resulting PLM index calculations when using as reference the latest clinical scoring guidelines. The agreement results are nevertheless comparable to the levels obtained in our work (ICC = 0.94), which use the recent WASM2016 standards [3]. Under this reference, our study is in fact the first one to set a reference for the agreement associated with manual derivation of the LM and PLM indices (ICC = 0.92 and 0.94, respectively). With regard to respiratory-derived indices, possibly the most widely reported is the AHI, with reliability scores for manual scoring nevertheless ranging widely between ICC 0.54-0.99 depending on the consulted study [5] [7] [47] [53]. Most likely, these differences are to a great extent driven by the specific rule used for the scoring of hypopneas. As stated before, it is widely accepted that agreement regarding scoring of hypopneas is lower as compared to that of apneas. This can also be observed by comparing ICC agreement values associated with AI and HI indices reported the literature. This is also the case in our study, with ICC agreement for the manual derivation of respiratory related indices showing a relatively high score for AHI (ICC = 0.98), following the expected trend of higher agreement for AI (ICC = 0.99) in comparison to HI (ICC = 0.60). In the case of the associated oxygen desaturation index, and ICC of 0.84 was obtained. Finally, and with regard to reliability of ArI indices resulting from manual scoring, literature shows high variability as well (ICC = 0.52-0.96 [45] [43] [7] [44] [5]). Our results for manual scoring approximately fit in the middle of that range (ICC = 0.68).

Some limitations of our study have to be mentioned as well. First, it is important to remark that absolute values of the various investigated performance scores are associated with one specific sleep lab. This study does not involve analysis of inter-scorer variability across multiple centers, and thus results might not generalize to other centers. In such scenario, the respective values of scoring agreement are expected to be lower in comparison due to the greater amount of variability involved [2] [47]. This study neither has attempted to quantify the corresponding levels of intra-scorer variability within any of the two examined approaches (manual or semi-automatic). Thus, it cannot completely be ruled-out that some of the differences between manual and semi-automatic approaches could be influenced by a component of intra-scorer variability effects, at the individual scorer case at least. Nevertheless, the relative high number of involved experts (12 in our study) should contribute to limit its the impact on the global results.

It should also be remarked that quality indicators derived from the semi-automatic scoring procedure are likely modulated by the reliability and performance of the specific automatic analysis algorithms used in the first instance. One might speculate with the idea that the better the algorithm, the higher the improvement on expert agreement with respect to the manual approach. However, there is no actual evidence that allows us to support this hypothesis. The usage of alternative automatic scoring methods might lead to different results. Regardless, our results support the hypothesis that semi-automatic algorithm can improve scoring quality in terms of both speed and resulting inter-scorer agreement. Also interesting, inter-scorer reliability studies available through literature, and this is no exception, implicitly assume that the outcome of all human scorers is equally valid. This might be a risky assumption, although there is no clear formula to discern who (out of a set of human experts) represents the best reference, and who does not. This propounds an interesting line of future research linked to another non-less interesting debate: can (full) automatic scoring outperform human experts? Of course, in terms of its capacity to correctly identify the relevant events associated with the physiological activity’s ground truth. There is no debate that automatic analysis can outperform manual scoring in terms of speed (and our study has shown this is also possible under a semi-automatic context). If, like in this case, the standard reference is subject to variability associated with human decision, it does not seem very plausible that any automatic algorithm could perform beyond the limit set by the average human agreement. After all, as stated before, deviations from such a reference do not necessarily correlate with the quality of the associated scorings. This is a subject that deserves more study.

Last but not least, one another possible limitation of this study relates to the number of PSGs involved in the evaluation of each analysis subtask. The relative high number of sleep experts involved partially counteracts this fact, and indeed, the number has proven enough to reach statistical significance among many of the reported hypothesis tests. However, a higher number of PSGs per task would be in general desirable. More specifically, for those cases in which the reported trends did not achieve significant effects, the question remains open on whether this could be attributed to the relative small PSG sample size. Notice, on the other hand, that post-hoc power analyses were consciously omitted because no useful conclusions are expected from them [54]. A higher sample size would also contribute to spread the bias risk due to demographic and physiological subject variability. Unfortunately, the chosen sample size was imposed by the available resources; thus this was not a design parameter we were able to tune. As noticed, scoring of PSG data is complex and time-consuming, and expert’s time is expensive and scant.

In conclusion, our results provide an updated reference for inter-scorer agreement levels and scoring times associated with both manual and semi-automatic scoring of PSG studies. We have systematically analyzed and compared the resulting differences, showing that the use of semi-automatic scoring can improve both speed and consistency of the PSG analysis outcomes. Benefits include reduction of the associated diagnostic costs, shortening of patient’s waiting lists, and overall improved productivity. In addition, enhancement of inter-scorer agreement leads to higher repeatability and quality of the diagnosis. More work has to be done to investigate generalization of these results by increasing the subject sample and its heterogeneity. Future work should also assess the effects of inter-center and intra-expert scoring variability, and goodness of fully automatic scoring in comparison to manual and semi-automatic approaches.

## Data Availability

All data produced in the present study are available upon reasonable request to the authors

## ACKNOWLEDGMENT

This study has been initiated at Haaglanden Medisch Centrum under project number 2019-073. The study has also been partially funded under project ED431H 2020/10 of Xunta de Galicia. Authors wish to acknowledge the support received from the Centro de Investigación de Galicia “CITIC”, funded by Xunta de Galicia and the European Union (European Regional Development Fund-Galicia 2014-2020 Program), by grant ED431G 2019/01.

