## Supplementary Materials for "Computer-assisted analysis of polysomnographic recordings improves inter-scorer associated agreement and scoring times"

**Corresponding author:**

**Supplementary materials**

Common interpretative considerations: respective distributions are characterized using the five-number summary as *p50 [minimum, p25, p75, maximum]*, where *XX* in the *pXX* notation refers to the corresponding percentile value. For further details, see Methods section in the main manuscript.

A. Scoring time. Individualized per-recording and per-scorer analyses

Table A1. Comparison of scoring time differences for sleep staging between manual and semi-automatic scoring approaches at the recording level

| **Sleep stating** | **n** | **Manual** | **Semi-Auto** | **Gain factor** | **Wilcoxon test *p*-value  (paired)** | **Effect size** |
| --- | --- | --- | --- | --- | --- | --- |
| SN1 | 12 | 40.59 [31.07, 34.72, 58.41, 142.63] | 29.09 [1.33, 19.32, 41.90, 147.76] | 1.40 | 0.0522 | 0.6021 |
| SN2 | 12 | 26.70 [14.91, 20.22, 39.03, 72.84] | 19.49 [5.14, 16.25, 27.93, 96.83] | 1.37 | 0.1294 | 0.3165 |
| SN3 | 12 | 57.14 [32.61, 45.02, 69.11, 166.32] | 48.05 [21.01, 32.51, 64.36, 180.04] | 1.19 | 0.2661 | 0.3616 |
| SN4 | 12 | 25.26 [18.50, 19.44, 34.39, 87.92] | 24.54 [14.31, 22.65, 31.01, 75.56] | 1.03 | 0.5693 | 0.2420 |
| SN5 | 12 | 16.97 [9.12, 13.83, 29.92, 47.88] | 12.86 [9.02, 11.42, 18.08, 36.70] | 1.32 | 0.0210* | 0.8328 |
| Overall | 60 | 32.62 [9.12, 21.74, 48.64, 166.32] | 24.54 [1.33, 16.25, 39.81, 180.04] | 1.33 | 0.0005* | 0.4297 |

**Statistically significant result*

Figure A1. Comparison of individual scorer times for sleep staging between manual and semi-automatic scoring approaches

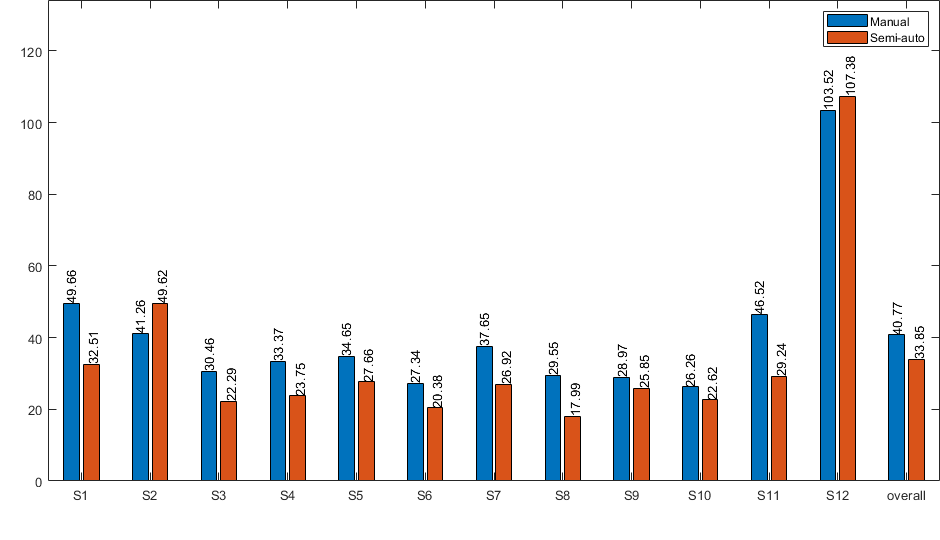

Table A2. Comparison of scoring time differences for detection of limb movement activity between manual and semi-automatic scoring approaches at the recording level

| **Limb movements** | **n** | **Manual** | **Semi-Auto** | **Gain factor** | **Wilcoxon test *p*-value  (paired)** | **Effect size** |
| --- | --- | --- | --- | --- | --- | --- |
| SN1 | 12 | 47.42 [25.22, 36.11, 65.35, 96.60] | 25.30 [12.16, 18.15, 28.31, 41.56] | 1.87 | 0.0005* | 1.5405 |
| SN2 | 12 | 27.13 [18.37, 20.16, 31.83, 40.69] | 11.40 [8.38, 9.00, 17.64, 31.55] | 2.38 | 0.0005* | 2.0332 |
| SN3 | 12 | 37.03 [23.76, 28.49, 45.91, 71.19] | 14.78 [9.74, 12.03, 17.30, 25.19] | 2.51 | 0.0005* | 2.0719 |
| SN4 | 12 | 68.79 [40.07, 60.49, 84.20, 126.08] | 20.36 [10.61, 15.54, 31.61, 55.67] | 3.38 | 0.0005* | 2.9294 |
| SN5 | 12 | 51.32 [28.04, 39.98, 65.97, 90.48] | 26.43 [12.37, 17.60, 38.51, 109.61] | 1.94 | 0.0210* | 0.9291 |
| Overall | 60 | 44.53 [18.37, 31.00, 65.30, 126.08] | 18.50 [8.38, 12.63, 26.73, 109.61] | 2.41 | < 0.0001* | 1.3475 |

**Statistically significant result*

Figure A2. Comparison of individual scorer times for detection of limb movement activity between manual and semi-automatic scoring approaches

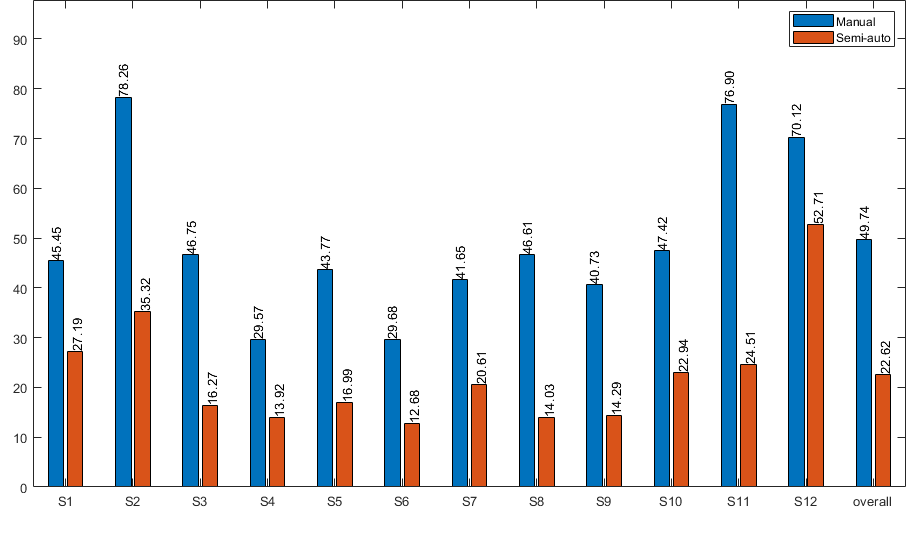

Table A3. Comparison of scoring time differences for analysis of respiratory activity between manual and semi-automatic scoring approaches at the recording level

| **Respiratory activity** | **n** | **Manual** | **Semi-Auto** | **Gain factor** | **Wilcoxon test *p*-value  (paired)** | **Effect size** |
| --- | --- | --- | --- | --- | --- | --- |
| SN1 | 12 | 18.02 [11.43, 16.97, 21.72, 43.78] | 11.26 [4.58, 8.68, 12.90, 41.26] | 1.60 | 0.0005* | 1.6927 |
| SN2 | 12 | 21.07 [10.79, 16.22, 32.51, 84.19] | 14.98 [5.90, 8.99, 20.34, 61.66] | 1.41 | 0.0342* | 0.7846 |
| SN3 | 12 | 56.56 [34.02, 51.09, 66.46, 119.81] | 23.50 [15.09, 18.11, 35.12, 74.93] | 2.41 | 0.0005* | 2.0932 |
| SN4 | 12 | 17.86 [9.59, 12.29, 27.44, 51.90] | 10.70 [4.87, 7.27, 13.04, 36.95] | 1.67 | 0.0005* | 1.3210 |
| SN5 | 12 | 27.26 [16.41, 20.28, 41.34, 104.06] | 17.19 [9.66, 14.04, 24.28, 55.81] | 1.59 | 0.0210* | 0.7914 |
| Overall | 60 | 23.81 [9.59, 17.62, 46.72, 119.81] | 14.58 [4.58, 10.46, 20.68, 74.93] | 1.63 | < 0.0001* | 0.9474 |

**Statistically significant result*

Figure A3. Comparison of individual scorer times for analysis of respiratory activity between manual and semi-automatic scoring approaches

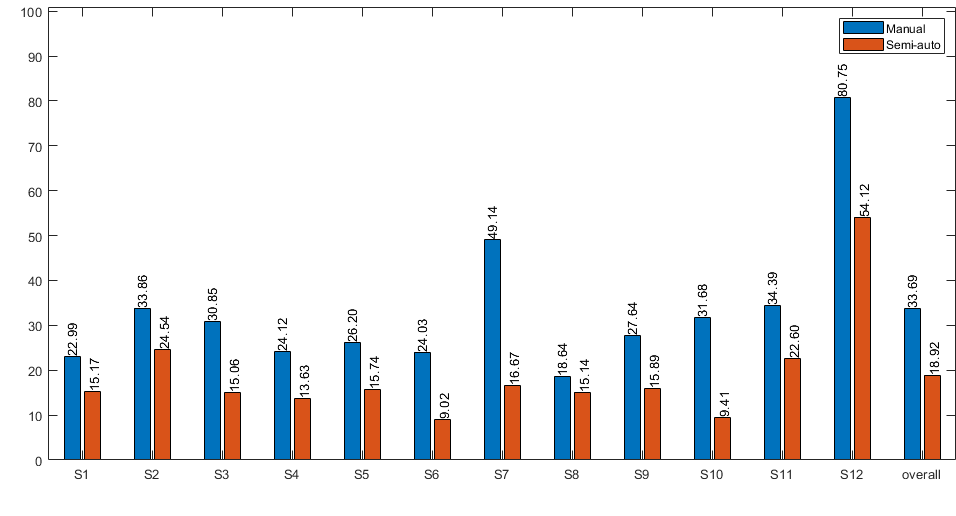

Table A4. Comparison of scoring time differences for EEG arousal detection between manual and semi-automatic scoring approaches at the recording level

| **EEG arousals** | **n** | **Manual** | **Semi-Auto** | **Gain factor** | **Wilcoxon test *p*-value  (paired)** | **Effect size** |
| --- | --- | --- | --- | --- | --- | --- |
| SN1 | 12 | 27.26 [16.59, 23.31, 40.51, 67.52] | 23.52 [15.33, 18.36, 26.70, 84.15] | 1.16 | 0.1294 | 0.5108 |
| SN2 | 12 | 20.66 [12.91, 17.77, 26.91, 48.25] | 17.49 [9.34, 12.84, 21.13, 66.78] | 1.18 | 0.1514 | 0.2718 |
| SN3 | 12 | 32.41 [18.75, 27.39, 38.13, 92.15] | 24.02 [5.06, 14.71, 37.49, 99.56] | 1.35 | 0.0425* | 0.7649 |
| SN4 | 12 | 27.81 [12.94, 17.40, 39.42, 69.77] | 19.55 [10.20, 15.26, 28.46, 91.92] | 1.42 | 0.2661 | 0.3343 |
| SN5 | 12 | 31.30 [20.05, 22.67, 42.37, 76.56] | 25.84 [12.27, 23.80, 35.22, 91.98] | 1.21 | 0.3013 | 0.2310 |
| Overall | 60 | 27.50 [12.91, 21.22, 37.65, 92.15] | 21.78 [5.06, 15.96, 28.59, 99.56] | 1.26 | 0.0011* | 0.4233 |

**Statistically significant result*

Figure A4. Comparison of individual scorer times for EEG arousal detection between manual and semi-automatic scoring approaches

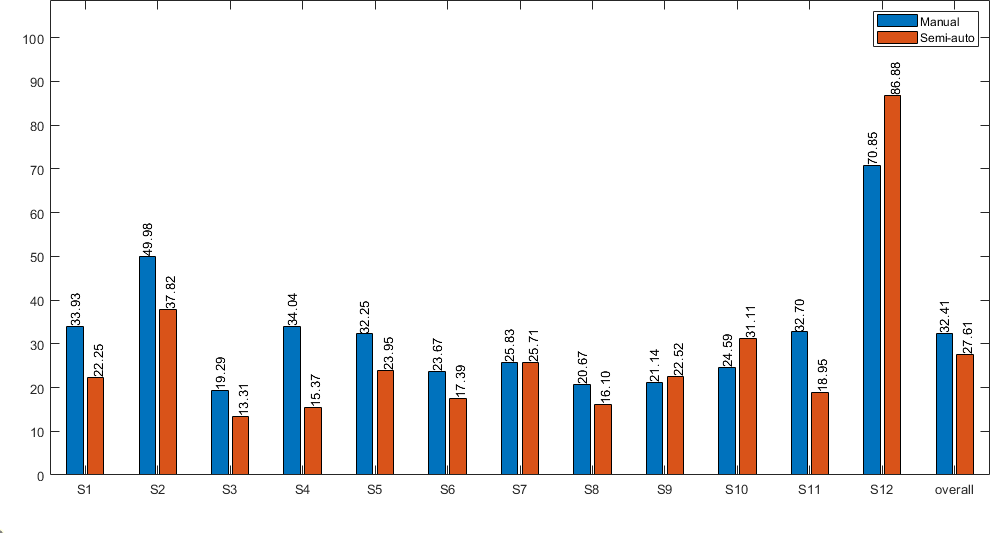

B. Kappa agreement. Individualized per-recording analyses

Table B1. Comparison of inter-scorer kappa agreement for the sleep stating scoring task between manual and semi-automatic scoring approaches at the recording level

| **Sleep stages (TIB)** | **n** | **Value a priori**** | **Paired agreement distribution summary descriptors** | | **Wilcoxon test *p*-value  (paired)** | **Effect size** |
| --- | --- | --- | --- | --- | --- | --- |
|  |  |  | Manual | Semi-Auto |  |  |
| SN1 | 66 | 50^th^ / 0.74 | 0.75 [0.47, 0.70, 0.80, 0.86] | 0.80 [0.70, 0.78, 0.82, 0.88] | < 0.0001* | -0.6600 |
| SN2 | 66 | 62.5^th^ / 0.77 | 0.66 [0.36, 0.62, 0.72, 0.83] | 0.77 [0.53, 0.72, 0.80, 0.93] | < 0.0001* | -1.2949 |
| SN3 | 66 | 12.5^th^ / 0.58 | 0.75 [0.52, 0.70, 0.78, 0.83] | 0.79 [0.64, 0.74, 0.82, 0.90] | < 0.0001* | -1.0604 |
| SN4 | 66 | 37.5^th^ / 0.70 | 0.77 [0.65, 0.74, 0.80, 0.83] | 0.82 [0.64, 0.78, 0.85, 0.94] | < 0.0001* | -0.5528 |
| SN5 | 66 | 87.5^th^ / 0.83 | 0.80 [0.62, 0.77, 0.84, 0.89] | 0.84 [0.69, 0.79, 0.87, 0.92] | 0.0015* | -0.4578 |
| Overall | 330 | --- | 0.76 [0.36, 0.69, 0.80, 0.89] | 0.80 [0.53, 0.76, 0.83, 0.94] | < 0.0001* | -0.7146 |

**Statistically significant result
**Value a priori: Distribution percentile and corresponding kappa agreement obtained during the selection procedure between the automatic algorithm and the retrospective expert scorings in the clinical pre-sample database (see Methods section)*

Table B2. Comparison of inter-scorer kappa agreement for the limb movements’ scoring task between manual and semi-automatic scoring approaches at the recording level

| **Limb movements (TIB)** | **n** | **Value a priori**** | **Paired agreement distribution summary descriptors** | | **Wilcoxon test *p*-value  (paired)** | **Effect size** |
| --- | --- | --- | --- | --- | --- | --- |
|  |  |  | Manual | Semi-Auto |  |  |
| SN1 | 66 | 50^th^ / 0.96 | 0.62 [0.46, 0.56, 0.69, 0.82] | 0.87 [0.77, 0.85, 0.93, 0.97] | < 0.0001* | -2.8314 |
| SN2 | 66 | 37.5^th^ / 0.95 | 0.71 [0.45, 0.64, 0.75, 0.87] | 0.91 [0.79, 0.87, 0.93, 0.98] | < 0.0001* | -2.5934 |
| SN3 | 66 | 62.5^th^ / 0.98 | 0.78 [0.57, 0.67, 0.84, 0.92] | 0.96 [0.79, 0.94, 0.97, 0.99] | < 0.0001* | -1.7358 |
| SN4 | 66 | 87.5^th^ / 1.00 | 0.79 [0.67, 0.75, 0.82, 0.91] | 0.94 [0.82, 0.91, 0.96, 0.98] | < 0.0001* | -2.5309 |
| SN5 | 66 | 12.5^th^ / 0.86 | 0.71 [0.47, 0.62, 0.77, 0.86] | 0.86 [0.78, 0.84, 0.89, 0.94] | < 0.0001* | -1.7866 |
| Overall | 330 | --- | 0.72 [0.45, 0.64, 0.79, 0.92] | 0.91 [0.77, 0.86, 0.95, 0.99] | < 0.0001* | -2.0223 |

**Statistically significant result
**Value a priori: Distribution percentile and corresponding kappa agreement obtained during the selection procedure between the automatic algorithm and the retrospective expert scorings in the clinical pre-sample database (see Methods section)*

Table B3. Comparison of inter-scorer kappa agreement for the limb movements’ scoring task in wakefulness between manual and semi-automatic scoring approaches at the recording level

| **Limb movements (Wake)** | **n** | **Paired agreement distribution summary descriptors** | | **Wilcoxon test *p*-value  (paired)** | **Effect size** |
| --- | --- | --- | --- | --- | --- |
|  |  | Manual | Semi-Auto |  |  |
| SN1 | 66 | 0.52 [0.28, 0.45, 0.60, 0.78] | 0.82 [0.66, 0.79, 0.91, 0.97] | < 0.0001* | -2.4747 |
| SN2 | 66 | 0.55 [0.19, 0.40, 0.65, 0.82] | 0.84 [0.66, 0.78, 0.89, 0.99] | < 0.0001* | -1.7473 |
| SN3 | 66 | 0.78 [0.57, 0.69, 0.83, 0.91] | 0.97 [0.78, 0.95, 0.98, 0.99] | < 0.0001* | -1.7562 |
| SN4 | 66 | 0.73 [0.54, 0.66, 0.79, 0.90] | 0.92 [0.69, 0.88, 0.96, 0.99] | < 0.0001* | -1.9417 |
| SN5 | 66 | 0.72 [0.51, 0.65, 0.78, 0.87] | 0.89 [0.79, 0.86, 0.92, 0.95] | < 0.0001* | -2.3142 |
| Overall | 330 | 0.67 [0.19, 0.57, 0.77, 0.91] | 0.89 [0.66, 0.82, 0.94, 0.99] | < 0.0001* | -1.7121 |

**Statistically significant result*

Table B4. Comparison of inter-scorer kappa agreement for the limb movements’ scoring task in sleep between manual and semi-automatic scoring approaches at the recording level

| **Limb movements (TST)** | **n** | **Paired agreement distribution summary descriptors** | | **Wilcoxon test *p*-value  (paired)** | **Effect size** |
| --- | --- | --- | --- | --- | --- |
|  |  | Manual | Semi-Auto |  |  |
| SN1 | 66 | 0.70 [0.49, 0.62, 0.75, 0.83] | 0.92 [0.80, 0.89, 0.94, 0.97] | < 0.0001* | -2.6375 |
| SN2 | 66 | 0.75 [0.49, 0.67, 0.80, 0.88] | 0.92 [0.80, 0.89, 0.94, 0.99] | < 0.0001* | -2.4712 |
| SN3 | 66 | 0.76 [0.53, 0.62, 0.83, 0.92] | 0.95 [0.77, 0.91, 0.96, 0.99] | < 0.0001* | -1.6278 |
| SN4 | 66 | 0.82 [0.73, 0.78, 0.85, 0.92] | 0.95 [0.88, 0.94, 0.96, 0.98] | < 0.0001* | -2.7504 |
| SN5 | 66 | 0.70 [0.39, 0.53, 0.77, 0.89] | 0.82 [0.66, 0.78, 0.85, 0.93] | < 0.0001* | -1.0254 |
| Overall | 330 | 0.75 [0.39, 0.65, 0.81, 0.92] | 0.92 [0.66, 0.86, 0.95, 0.99] | < 0.0001* | -1.6704 |

**Statistically significant result*

Table B5. Comparison of inter-scorer kappa agreement for the respiratory analysis scoring task between manual and semi-automatic scoring approaches at the recording level

| **Respiratory Events (Apnea, Hypopnea, RERA)  (TIB)** | **n** | **Value a priori**** | **Paired agreement distribution summary descriptors** | | **Wilcoxon test *p*-value  (paired)** | **Effect size** |
| --- | --- | --- | --- | --- | --- | --- |
|  |  |  | Manual | Semi-Auto |  |  |
| SN1 | 66 | 62.5^th^ / 0.86 | 0.75 [0.66, 0.71, 0.78, 0.86] | 0.86 [0.75, 0.84, 0.89, 0.94] | < 0.0001* | -1.7838 |
| SN2 | 66 | 12.5^th^ / 0.37 | 0.47 [0.22, 0.34, 0.52, 0.68] | 0.51 [0.30, 0.43, 0.60, 0.77] | 0.0021* | -0.4249 |
| SN3 | 66 | 87.5^th^ / 0.88 | 0.85 [0.72, 0.82, 0.87, 0.92] | 0.92 [0.88, 0.91, 0.94, 0.95] | < 0.0001* | -2.3013 |
| SN4 | 66 | 37.5^th^ / 0.65 | 0.46 [0.06, 0.37, 0.53, 0.68] | 0.47 [0.20, 0.40, 0.59, 0.78] | 0.0019* | -0.4262 |
| SN5 | 66 | 50^th^ / 0.68 | 0.47 [0.18, 0.38, 0.53, 0.62] | 0.64 [0.47, 0.59, 0.68, 0.80] | < 0.0001* | -1.9555 |
| Overall | 330 | --- | 0.55 [0.06, 0.43, 0.78, 0.92] | 0.66 [0.20, 0.53, 0.89, 0.95] | < 0.0001* | -0.8315 |

**Statistically significant result
**Value a priori: Distribution percentile and corresponding kappa agreement obtained during the selection procedure between the automatic algorithm and the retrospective expert scorings in the clinical pre-sample database (see Methods section)*

Table B6. Comparison of inter-scorer kappa agreement for the respiratory analysis scoring task (apneas only) between manual and semi-automatic scoring approaches at the recording level

| **Respiratory Events (Apneas only)  (TIB)** | **n** | **Paired agreement distribution summary descriptors** | | **Wilcoxon test *p*-value  (paired)** | **Effect size** |
| --- | --- | --- | --- | --- | --- |
|  |  | Manual | Semi-Auto |  |  |
| SN1 | 66 | 0.75 [0.56, 0.70, 0.79, 0.92] | 0.85 [0.72, 0.82, 0.94, 1.00] | < 0.0001* | -1.0425 |
| SN2 | 66 | 0.33 [-0.00, 0.00, 0.60, 0.77] | 0.00 [-0.00, 0.00, 0.41, 1.00] | 0.2374 | 0.1293 |
| SN3 | 66 | 0.86 [0.75, 0.84, 0.88, 0.93] | 0.96 [0.91, 0.95, 0.96, 0.98] | < 0.0001* | -2.7070 |
| SN4 | 66 | 1.00 [0.00, 1.00, 1.00, 1.00] | 1.00 [1.00, 1.00, 1.00, 1.00] | 0.0010* | -0.4438 |
| SN5 | 66 | 0.32 [-0.00, 0.23, 0.44, 0.65] | 0.58 [0.33, 0.48, 0.65, 0.82] | < 0.0001* | -1.1821 |
| Overall | 330 | 0.74 [-0.00, 0.35, 0.88, 1.00] | 0.88 [-0.00, 0.57, 0.98, 1.00] | < 0.0001* | -0.3783 |

**Statistically significant result*

Table B7. Comparison of inter-scorer kappa agreement for the respiratory analysis scoring task (hypopneas only) between manual and semi-automatic scoring approaches at the recording level

| **Respiratory Events (Hypopneas only)  (TIB)** | **n** | **Paired agreement distribution summary descriptors** | | **Wilcoxon test *p*-value  (paired)** | **Effect size** |
| --- | --- | --- | --- | --- | --- |
|  |  | Manual | Semi-Auto |  |  |
| SN1 | 66 | 0.51 [0.29, 0.42, 0.59, 0.80] | 0.75 [0.59, 0.71, 0.81, 0.90] | < 0.0001* | -1.7069 |
| SN2 | 66 | 0.44 [0.16, 0.30, 0.53, 0.71] | 0.52 [0.31, 0.44, 0.62, 0.79] | 0.0001* | -0.5570 |
| SN3 | 66 | 0.44 [0.15, 0.35, 0.52, 0.72] | 0.58 [0.38, 0.55, 0.63, 0.72] | < 0.0001* | -1.1295 |
| SN4 | 66 | 0.46 [0.06, 0.37, 0.54, 0.68] | 0.47 [0.20, 0.40, 0.59, 0.78] | 0.0021* | -0.4231 |
| SN5 | 66 | 0.46 [0.14, 0.34, 0.52, 0.60] | 0.63 [0.48, 0.59, 0.67, 0.81] | < 0.0001* | -1.8539 |
| Overall | 330 | 0.46 [0.06, 0.36, 0.53, 0.80] | 0.61 [0.20, 0.51, 0.68, 0.90] | < 0.0001* | -0.9569 |

**Statistically significant result*

Table B8. Comparison of inter-scorer kappa agreement for the EEG arousals’ scoring task between manual and semi-automatic scoring approaches at the recording level

| **EEG Arousals (TIB)** | **n** | **Value a priori**** | **Paired agreement distribution summary descriptors** | | **Wilcoxon test *p*-value  (paired)** | **Effect size** |
| --- | --- | --- | --- | --- | --- | --- |
|  |  |  | Manual | Semi-Auto |  |  |
| SN1 | 66 | 12.5^th^ / 0.30 | 0.54 [0.40, 0.48, 0.59, 0.66] | 0.54 [0.38, 0.50, 0.57, 0.65] | 0.6944 | 0.0360 |
| SN2 | 66 | 37.5^th^ / 0.51 | 0.48 [0.19, 0.41, 0.54, 0.67] | 0.56 [0.41, 0.52, 0.62, 0.67] | < 0.0001* | -1.0501 |
| SN3 | 66 | 50^th^ / 0.56 | 0.60 [0.38, 0.56, 0.64, 0.71] | 0.66 [0.50, 0.62, 0.69, 0.87] | < 0.0001* | -0.8658 |
| SN4 | 66 | 67.5^th^ / 0.62 | 0.67 [0.38, 0.62, 0.72, 0.80] | 0.73 [0.59, 0.67, 0.76, 0.82] | 0.0005* | -0.5154 |
| SN5 | 66 | 87.5^th^ - 0.81 | 0.64 [0.33, 0.52, 0.70, 0.79] | 0.71 [0.57, 0.69, 0.74, 0.81] | < 0.0001* | -0.7811 |
| Overall | 330 | --- | 0.58 [0.19, 0.48, 0.65, 0.80] | 0.65 [0.38, 0.56, 0.71, 0.87] | < 0.0001* | -0.6166 |

**Statistically significant result
**Value a priori: Distribution percentile and corresponding kappa agreement obtained during the selection procedure between the automatic algorithm and the retrospective expert scorings in the clinical pre-sample database (see Methods section)*

C. Diagnostic indices. Per-recording analyses of associated inter-scorer variability

Table C1. Comparison of derived diagnostic indices for Sleep Efficiency between manual and semi-automatic scoring approaches at the recording level

| **Sleep Efficiency (SE)** | **n** | **Summary of distributions** | | **Wilcoxon test *p*-value  (paired)** | **Effect size** | **Normalized standard deviation** | | **Brown-Forsythe *p*-value (unpaired)** | **Variance Ratio**** |
| --- | --- | --- | --- | --- | --- | --- | --- | --- | --- |
|  |  | Manual | Semi-Auto |  |  | Manual | Semi-auto |  |  |
| SN1 | 12 | 94.75 [87.00, 93.19, 95.19, 96.37] | 93.75 [92.57, 92.91, 94.86, 96.03] | 0.7485 | -0.0293 | 0.03 | 0.01 | 0.4720 | 4.2720 |
| SN2 | 12 | 95.82 [93.48, 95.41, 96.78, 98.03] | 96.51 [95.54, 95.89, 97.13, 97.61] | 0.1133 | -0.4941 | 0.01 | 0.01 | 0.0925 | 3.6102 |
| SN3 | 12 | 89.45 [87.56, 88.23, 90.22, 92.26] | 90.31 [89.63, 89.82, 90.73, 91.29] | 0.0771 | -0.5589 | 0.02 | 0.01 | 0.0099* | 7.6163 |
| SN4 | 12 | 69.29 [66.17, 68.05, 71.90, 74.08] | 71.12 [67.01, 70.59, 71.90, 76.37] | 0.0269* | -0.6550 | 0.04 | 0.03 | 0.1596 | 1.6385 |
| SN5 | 12 | 24.09 [22.79, 23.24, 25.61, 34.41] | 26.45 [23.24, 25.55, 27.81, 31.25] | 0.0791 | -0.4044 | 0.13 | 0.08 | 0.6156 | 2.5403 |
| Overall | 60 | 89.11 [22.79, 68.05, 94.87, 98.03] | 90.31 [23.24, 70.59, 94.86, 97.61] | 0.0004* | -0.3717 | 0.06 | 0.04 | 0.1384 | 2.5018 |

**Statistically significant result
**Calculated as Var[Manual]/Var[Semi-Auto], thus values > 1 point out toward higher dispersion (variability) among the manual scoring distribution*

Table C2. Comparison of derived diagnostic indices for Sleep Onset Latency between manual and semi-automatic scoring approaches at the recording level

| **Sleep Onset Latency (SOL)** | **n** | **Summary of distributions** | | **Wilcoxon test *p*-value  (paired)** | **Effect size** | **Normalized standard deviation** | | **Brown-Forsythe *p*-value (unpaired)** | **Variance Ratio**** |
| --- | --- | --- | --- | --- | --- | --- | --- | --- | --- |
|  |  | Manual | Semi-Auto |  |  | Manual | Semi-auto |  |  |
| SN1 | 12 | 0.63 [0.63, 0.63, 3.38, 5.13] | 1.88 [0.13, 0.63, 2.88, 11.13] | 0.5898 | -0.2672 | 0.92 | 1.20 | 0.9128 | 0.5898 |
| SN2 | 12 | 3.00 [1.50, 2.75, 5.00, 5.50] | 2.50 [2.50, 2.50, 3.00, 4.50] | 0.1367 | 0.5499 | 0.37 | 0.20 | 0.1675 | 3.2171 |
| SN3 | 12 | 3.21 [0.00, 0.71, 6.71, 7.21] | 1.96 [0.00, 0.71, 6.21, 6.21] | 0.5342 | 0.1651 | 0.81 | 0.93 | 0.5356 | 0.7597 |
| SN4 | 12 | 106.91 [5.41, 15.66, 107.41, 107.41] | 106.41 [1.91, 16.91, 106.91, 107.41] | 0.5938 | -0.0526 | 0.63 | 0.62 | 0.9670 | 1.0401 |
| SN5 | 12 | 168.20 [73.20, 148.45, 169.70, 169.70] | 112.95 [103.20, 103.20, 168.20, 169.70] | 0.1230 | 0.5270 | 0.20 | 0.24 | 0.3818 | 0.7176 |
| Overall | 60 | 5.27 [0.00, 2.50, 107.41, 169.70] | 4.11 [0.00, 2.31, 104.81, 169.70] | 0.0629 | 0.2227 | 0.62 | 0.72 | 0.8598 | 0.7455 |

**Statistically significant result
**Calculated as Var[Manual]/Var[Semi-Auto], thus values > 1 point out toward higher dispersion (variability) among the manual scoring distribution*

Table C3. Comparison of derived diagnostic indices for Wake After Sleep Onset between manual and semi-automatic scoring approaches at the recording level

| **Wake After Sleep Onset (WASO)** | **n** | **Summary of distributions** | | **Wilcoxon test *p*-value  (paired)** | **Effect size** | **Normalized standard deviation** | | **Brown-Forsythe *p*-value (unpaired)** | **Variance Ratio**** |
| --- | --- | --- | --- | --- | --- | --- | --- | --- | --- |
|  |  | Manual | Semi-Auto |  |  | Manual | Semi-auto |  |  |
| SN1 | 12 | 23.53 [16.28, 21.52, 30.46, 58.28] | 28.02 [17.78, 23.02, 31.75, 33.24] | 0.7334 | 0.0303* | 0.40 | 0.19 | 0.4846 | 4.1972 |
| SN2 | 12 | 15.09 [7.12, 11.59, 16.61, 23.58] | 12.61 [8.62, 10.36, 14.84, 16.12] | 0.1050 | 0.4941 | 0.31 | 0.19 | 0.1804 | 2.6980 |
| SN3 | 12 | 86.20 [63.25, 79.95, 96.15, 101.74] | 79.18 [71.25, 75.70, 83.23, 84.76] | 0.0771 | 0.5588 | 0.14 | 0.06 | 0.0152* | 6.3587 |
| SN4 | 12 | 145.76 [124.46, 134.84, 151.77, 160.76] | 137.03 [113.31, 134.03, 139.51, 156.76] | 0.0269 | 0.6577 | 0.08 | 0.07 | 0.2223 | 1.3734 |
| SN5 | 12 | 333.66 [289.49, 327.69, 337.39, 339.39] | 323.62 [302.99, 318.24, 327.62, 337.38] | 0.0640 | 0.3964 | 0.04 | 0.03 | 0.6838 | 2.2134 |
| Overall | 60 | 86.20 [7.12, 22.27, 151.77, 339.39] | 79.18 [8.62, 23.02, 139.51, 337.38] | 0.0003 | 0.3929 | 0.23 | 0.12 | 0.0307* | 3.4201 |

**Statistically significant result
**Calculated as Var[Manual]/Var[Semi-Auto], thus values > 1 point out toward higher dispersion (variability) among the manual scoring distribution*

Table C4. Comparison of derived diagnostic indices for Limb Movement Index (LMI) between manual and semi-automatic scoring approaches at the recording level

| **Limb Movement Index (LMI)** | **n** | **Summary of distributions** | | **Wilcoxon test *p*-value  (paired)** | **Effect size** | **Normalized standard deviation** | | **Brown-Forsythe *p*-value (unpaired)** | **Variance Ratio**** |
| --- | --- | --- | --- | --- | --- | --- | --- | --- | --- |
|  |  | Manual | Semi-Auto |  |  | Manual | Semi-auto |  |  |
| SN1 | 12 | 25.94 [23.01, 23.54, 30.98, 35.79] | 31.73 [29.62, 30.30, 32.48, 35.19] | 0.0122* | -0.9204 | 0.17 | 0.05 | 0.0164* | 11.0770 |
| SN2 | 12 | 19.78 [14.28, 17.92, 22.38, 25.28] | 21.27 [20.52, 20.89, 22.46, 23.65] | 0.0732 | -0.6051 | 0.16 | 0.05 | 0.0098* | 12.3865 |
| SN3 | 12 | 17.64 [14.87, 15.61, 21.06, 22.02] | 19.18 [17.21, 17.93, 19.68, 20.27] | 0.2915 | -0.2779 | 0.15 | 0.05 | 0.0013* | 8.2051 |
| SN4 | 12 | 104.81 [91.66, 101.25, 111.77, 116.78] | 107.69 [100.32, 105.32, 108.97, 115.25] | 0.5059 | -0.2701 | 0.08 | 0.03 | 0.0317* | 4.8308 |
| SN5 | 12 | 59.52 [16.24, 40.61, 70.19, 81.94] | 50.42 [32.00, 40.48, 61.82, 67.88] | 0.4316 | 0.1816 | 0.39 | 0.24 | 0.2112 | 2.6541 |
| Overall | 60 | 25.27 [14.28, 19.78, 70.19, 116.78] | 31.73 [17.21, 20.89, 61.82, 115.25] | 0.0999 | -0.0763 | 0.21 | 0.11 | 0.0003* | 3.5592 |

**Statistically significant result
**Calculated as Var[Manual]/Var[Semi-Auto], thus values > 1 point out toward higher dispersion (variability) among the manual scoring distribution*

Table C5. Comparison of derived diagnostic indices for Periodic Limb Movement Index (PLMI) between manual and semi-automatic scoring approaches at the recording level

| **Periodic Limb Movement Index (PLMI)** | **n** | **Summary of distributions** | | **Wilcoxon test *p*-value  (paired)** | **Effect size** | **Normalized standard deviation** | | **Brown-Forsythe *p*-value (unpaired)** | **Variance Ratio**** |
| --- | --- | --- | --- | --- | --- | --- | --- | --- | --- |
|  |  | Manual | Semi-Auto |  |  | Manual | Semi-auto |  |  |
| SN1 | 12 | 12.41 [8.57, 10.16, 15.11, 20.60] | 13.84 [12.03, 13.46, 14.52, 16.84] | 0.2036 | -0.2777 | 0.30 | 0.10 | 0.0038* | 9.1267 |
| SN2 | 12 | 12.79 [6.99, 10.41, 13.75, 14.72] | 13.32 [12.49, 12.94, 13.46, 15.32] | 0.0771 | -0.6003 | 0.21 | 0.05 | 0.0052* | 15.5409 |
| SN3 | 12 | 1.46 [0.58, 0.87, 2.77, 4.37] | 2.48 [1.75, 2.41, 3.28, 4.08] | 0.0791 | -0.6395 | 0.65 | 0.23 | 0.0144* | 7.7557 |
| SN4 | 12 | 93.70 [83.17, 91.49, 99.72, 103.37] | 96.92 [89.11, 95.98, 97.94, 102.01] | 0.3203 | -0.3522 | 0.06 | 0.03 | 0.0589 | 3.7146 |
| SN5 | 12 | 52.97 [11.39, 31.16, 61.70, 71.27] | 42.06 [22.30, 32.12, 53.70, 61.09] | 0.3013 | 0.1796 | 0.45 | 0.29 | 0.3426 | 2.3274 |
| Overall | 60 | 13.91 [0.58, 9.20, 61.70, 103.37] | 14.14 [1.75, 12.79, 53.70, 102.01] | 0.1842 | -0.0245 | 0.38 | 0.11 | < 0.0001* | 4.9196 |

**Statistically significant result
**Calculated as Var[Manual]/Var[Semi-Auto], thus values > 1 point out toward higher dispersion (variability) among the manual scoring distribution*

Table C6. Comparison of derived diagnostic indices for Apnea-Hypopnea Index (AHI) between manual and semi-automatic scoring approaches at the recording level

| **Apnea-Hypopnea Index (AHI)** | **n** | **Summary of distributions** | | **Wilcoxon test *p*-value  (paired)** | **Effect size** | **Normalized standard deviation** | | **Brown-Forsythe *p*-value (unpaired)** | **Variance Ratio**** |
| --- | --- | --- | --- | --- | --- | --- | --- | --- | --- |
|  |  | Manual | Semi-Auto |  |  | Manual | Semi-auto |  |  |
| SN1 | 12 | 5.96 [3.97, 5.09, 6.39, 6.56] | 6.21 [3.45, 5.53, 6.56, 6.91] | 0.2915 | -0.1499 | 0.15 | 0.18 | 0.9174 | 0.7009 |
| SN2 | 12 | 3.71 [0.21, 1.75, 5.67, 6.60] | 4.74 [1.03, 4.54, 6.19, 7.84] | 0.2256 | -0.3764 | 0.56 | 0.39 | 0.1495 | 1.9995 |
| SN3 | 12 | 53.73 [44.73, 51.75, 54.31, 55.79] | 56.20 [53.81, 55.38, 57.03, 58.10] | 0.0010* | -1.0041 | 0.05 | 0.02 | 0.3391 | 5.1273 |
| SN4 | 12 | 3.49 [0.17, 2.58, 4.08, 5.65] | 4.49 [1.99, 2.66, 5.49, 7.15] | 0.1685 | -0.4898 | 0.43 | 0.40 | 0.8031 | 1.1386 |
| SN5 | 12 | 9.47 [3.56, 6.20, 12.47, 14.25] | 13.32 [7.98, 10.54, 14.61, 17.67] | 0.0098* | -0.9169 | 0.37 | 0.24 | 0.0945 | 2.4070 |
| Overall | 60 | 5.86 [0.17, 3.71, 12.47, 55.79] | 6.54 [1.03, 4.70, 14.61, 58.10] | < 0.0001* | -0.0906 | 0.35 | 0.28 | 0.2058 | 1.6157 |

**Statistically significant result
**Calculated as Var[Manual]/Var[Semi-Auto], thus values > 1 point out toward higher dispersion (variability) among the manual scoring distribution*

Table C7. Comparison of derived diagnostic indices for Apnea Index (AI) between manual and semi-automatic scoring approaches at the recording level

| **Apnea Index (AI)** | **n** | **Summary of distributions** | | **Wilcoxon test *p*-value  (paired)** | **Effect size** | **Normalized standard deviation** | | **Brown-Forsythe *p*-value (unpaired)** | **Variance Ratio**** |
| --- | --- | --- | --- | --- | --- | --- | --- | --- | --- |
|  |  | Manual | Semi-Auto |  |  | Manual | Semi-auto |  |  |
| SN1 | 12 | 3.11 [2.24, 2.76, 3.46, 5.01] | 3.02 [2.59, 2.76, 3.54, 3.80] | 0.6797 | 0.1934 | 0.23 | 0.13 | 0.4082 | 3.1926 |
| SN2 | 12 | 0.72 [0.00, 0.31, 1.03, 1.44] | 0.41 [0.00, 0.00, 0.62, 1.24] | 0.2500 | 0.4379 | 0.63 | 1.07 | 0.1241 | 0.3393 |
| SN3 | 12 | 50.59 [42.75, 49.11, 51.83, 52.49] | 51.50 [48.86, 50.09, 53.14, 53.48] | 0.0332* | -0.6133 | 0.06 | 0.03 | 0.4382 | 2.8018 |
| SN4 | 12 | 0.00 [0.00, 0.00, 0.00, 0.17] | 0.00 [0.00, 0.00, 0.00, 0.00] | 1.0000 | 0.2887 | 3.46 | 0.00 | 0.3282 | --- |
| SN5 | 12 | 0.92 [0.43, 0.50, 1.57, 2.71] | 1.57 [0.86, 1.43, 2.07, 2.71] | 0.0439* | -0.7011 | 0.66 | 0.34 | 0.0423* | 3.6581 |
| Overall | 60 | 1.26 [0.00, 0.31, 3.46, 52.49] | 1.57 [0.00, 0.00, 3.54, 53.48] | 0.0840 | -0.2483 | 1.55 | 0.64 | 0.3685 | 5.9561 |

**Statistically significant result
**Calculated as Var[Manual]/Var[Semi-Auto], thus values > 1 point out toward higher dispersion (variability) among the manual scoring distribution*

Table C8. Comparison of derived diagnostic indices for Hypopnea Index (HI) between manual and semi-automatic scoring approaches at the recording level

| **Hypopnea Index (HI)** | **n** | **Summary of distributions** | | **Wilcoxon test *p*-value  (paired)** | **Effect size** | **Normalized standard deviation** | | **Brown-Forsythe *p*-value (unpaired)** | **Variance Ratio**** |
| --- | --- | --- | --- | --- | --- | --- | --- | --- | --- |
|  |  | Manual | Semi-Auto |  |  | Manual | Semi-auto |  |  |
| SN1 | 12 | 2.42 [0.69, 2.16, 2.93, 3.28] | 2.76 [0.35, 2.42, 3.62, 4.14] | 0.2729 | -0.2322 | 0.32 | 0.44 | 0.3838 | 0.5168 |
| SN2 | 12 | 2.99 [0.21, 1.23, 4.75, 5.16] | 4.54 [1.03, 3.92, 5.57, 7.22] | 0.1050 | -0.5410 | 0.61 | 0.40 | 0.0563 | 2.3194 |
| SN3 | 12 | 2.73 [1.82, 2.31, 3.14, 4.79] | 4.71 [2.31, 3.71, 5.70, 6.77] | 0.0024* | -1.2402 | 0.29 | 0.28 | 0.7957 | 1.1134 |
| SN4 | 12 | 3.41 [0.17, 2.58, 4.08, 5.65] | 4.49 [1.99, 2.66, 5.49, 7.15] | 0.1685 | -0.4933 | 0.43 | 0.40 | 0.8395 | 1.1477 |
| SN5 | 12 | 7.70 [2.28, 5.78, 11.33, 13.54] | 11.12 [7.13, 8.91, 13.12, 15.68] | 0.0093* | -0.8548 | 0.43 | 0.25 | 0.0791 | 2.8708 |
| Overall | 60 | 3.10 [0.17, 2.38, 4.87, 13.54] | 4.55 [0.35, 3.13, 6.68, 15.68] | < 0.0001* | -0.6271 | 0.42 | 0.35 | 0.2702 | 1.4050 |

**Statistically significant result
**Calculated as Var[Manual]/Var[Semi-Auto], thus values > 1 point out toward higher dispersion (variability) among the manual scoring distribution*

Table C9. Comparison of derived diagnostic indices for Oxygen Desaturation Index (ODI) between manual and semi-automatic scoring approaches at the recording level

| **Oxygen Desaturation Index (ODI)** | **n** | **Summary of distributions** | | **Wilcoxon test *p*-value  (paired)** | **Effect size** | **Normalized standard deviation** | | **Brown-Forsythe *p*-value (unpaired)** | **Variance Ratio**** |
| --- | --- | --- | --- | --- | --- | --- | --- | --- | --- |
|  |  | Manual | Semi-Auto |  |  | Manual | Semi-auto |  |  |
| SN1 | 12 | 6.04 [3.28, 5.00, 6.47, 6.91] | 6.13 [5.35, 5.62, 6.56, 7.25] | 0.1563 | -0.5125 | 0.20 | 0.11 | 0.0978 | 3.6811 |
| SN2 | 12 | 10.41 [2.47, 5.36, 12.58, 14.64] | 12.68 [7.42, 10.62, 13.61, 15.05] | 0.0220* | 0.7914 | 0.47 | 0.19 | 0.0053* | 6.0152 |
| SN3 | 12 | 49.02 [3.14, 45.97, 64.37, 72.30] | 64.62 [49.19, 64.21, 65.94, 68.50] | 0.0186* | -0.6892 | 0.36 | 0.10 | 0.0296* | 13.8934 |
| SN4 | 12 | 3.49 [0.33, 2.41, 4.24, 4.99] | 5.90 [3.49, 5.16, 5.98, 6.48] | 0.0024* | -1.2408 | 0.42 | 0.14 | 0.0097* | 8.6081 |
| SN5 | 12 | 12.18 [3.71, 7.70, 14.68, 16.25] | 14.89 [10.55, 14.11, 15.89, 17.81] | 0.0171* | -0.8690 | 0.38 | 0.13 | 0.0127* | 8.5842 |
| Overall | 60 | 6.48 [0.33, 4.24, 14.38, 72.30] | 12.04 [3.49, 6.01, 15.89, 68.50] | < 0.0001* | -0.4759 | 0.36 | 0.13 | < 0.0001* | 7.5395 |

**Statistically significant result
**Calculated as Var[Manual]/Var[Semi-Auto], thus values > 1 point out toward higher dispersion (variability) among the manual scoring distribution*

Table C10. Comparison of derived diagnostic indices for Arousal Index (ArI) between manual and semi-automatic scoring approaches at the recording level

| **Arousal Index (ArI)** | **n** | **Summary of distributions** | | **Wilcoxon test *p*-value  (paired)** | **Effect size** | **Normalized standard deviation** | | **Brown-Forsythe *p*-value (unpaired)** | **Variance Ratio**** |
| --- | --- | --- | --- | --- | --- | --- | --- | --- | --- |
|  |  | Manual | Semi-Auto |  |  | Manual | Semi-auto |  |  |
| SN1 | 12 | 24.24 [15.63, 19.43, 26.84, 36.06] | 22.27 [13.60, 17.83, 25.52, 32.12] | 0.2661 | 0.3671 | 0.27 | 0.24 | 0.7511 | 1.3309 |
| SN2 | 12 | 8.51 [3.84, 6.13, 11.52, 17.20] | 9.60 [5.38, 7.53, 11.21, 12.20] | 0.7910 | -0.1051 | 0.43 | 0.24 | 0.0830 | 3.2394 |
| SN3 | 12 | 18.01 [12.98, 15.85, 24.25, 29.78] | 19.18 [13.50, 17.48, 24.10, 32.56] | 0.7910 | -0.2315 | 0.28 | 0.26 | 0.8114 | 1.1390 |
| SN4 | 12 | 14.29 [10.12, 11.70, 15.32, 18.87] | 15.07 [12.95, 14.12, 17.87, 23.32] | 0.0063* | -0.9293 | 0.20 | 0.20 | 0.9421 | 0.9210 |
| SN5 | 12 | 27.70 [15.12, 24.61, 32.30, 35.33] | 31.05 [23.70, 26.97, 33.71, 36.37] | 0.0425* | -0.5423 | 0.20 | 0.14 | 0.4315 | 2.0931 |
| Overall | 60 | 17.57 [3.84, 12.98, 25.64, 36.06] | 18.38 [5.38, 13.82, 25.41, 36.37] | 0.1756 | -0.1866 | 0.28 | 0.21 | 0.1300 | 1.7129 |

**Statistically significant result
**Calculated as Var[Manual]/Var[Semi-Auto], thus values > 1 point out toward higher dispersion (variability) among the manual scoring distribution*

D. Correlation analyses for automatic selection of PSG recordings involved in the study

The main text manuscript describes an automatic PSG selection procedure to avoid selection bias. There the hypothesis is stated that automatic scoring difficulty would be correlated with manual scoring difficulty: the lower (higher) the agreement between the automatic and the clinical reference, the lower (higher) the expected inter-rater agreement should be.

Table D1 shows the results of correlation analyses between the automatic-clinical kappa agreement scores obtained during the selection process, and the respective levels of inter-scorer agreement obtained after the rescoring process has been performed. Individual a priori and a posteriori agreement kappa’s involved in the analyses can be found in the corresponding tables in section B of the Supplementary Materials.

Table D1. Correlation between automatic-clinical selection agreement with respect to manual and semi-automatic achieved agreements during rescoring tasks (using the kappa statistic)

| **Scoring task** | **n** | **Selection vs Manual** | | **Selection vs Semi-auto** | |
| --- | --- | --- | --- | --- | --- |
|  |  | *r* | *p*-value | *r* | *p*-value |
| Sleep staging | 5 | 0.0549 | 0.9301 | 0.3601 | 0.5516 |
| Limb Movements | 5 | 0.4423 | 0.4557 | 0.8591 | 0.0621 |
| Respiratory events | 5 | 0.7946 | 0.1083 | 0.8590 | 0.0622 |
| EEG Arousals | 5 | 0.5947 | 0.2902 | 0.8752 | 0.0519 |
| Altogether | 20 | 0.6645 | 0.0014* | 0.8789 | < 0.0001* |

**Statistically significant result*

Data from Table D1 shows heterogeneous individual per-task correlations that do not reach statistically significant levels. This is possibly due to the low number of cases involved per task in the corresponding calculations (only 5). When aggregating data from all the scoring tasks together, however, significant correlation effects are noticed. This is likely due to the increased number of available data points (n=20), therefore supporting the selection hypothesis. In general, higher correlation values are obtained in the context of the semi-automatic scoring scenario (r = 0.8789 vs r = 0.6645 with respect to the manual rescoring approach). This result is partially expected, as scorings derived from the semi-automatic approach more likely resemble the output of the pure automatic scoring algorithm (no human intervention) used during the selection process.
